# Age-structured non-pharmaceutical interventions for optimal control of COVID-19 epidemic

**DOI:** 10.1101/2020.06.23.20138099

**Authors:** Quentin Richard, Samuel Alizon, Marc Choisy, Mircea T. Sofonea, Ramsès Djidjou-Demasse

## Abstract

In an epidemic, individuals can widely differ in the way they spread the infection depending on their age or on the number of days they have been infected for. In the absence of pharmaceutical interventions such as a vaccine or treatment, non-pharmaceutical interventions (*e.g*. social distancing) are essential to mitigate the pandemic. We develop an original approach to identify the optimal age-stratified control strategy to implement as a function of the time since the onset of the epidemic. This is based on a model with a double continuous structure in terms of host age and time since infection. By applying optimal control theory to this model, we identify a solution that minimizes deaths and costs associated with the implementation of the control strategy itself. We also implement this strategy to three countries with contrasted age distributions (Burkina-Faso, France, and Vietnam). Overall, the optimal strategy varies over the course of the epidemic, with a more intense control early on, and depending on host age, with a stronger control for the older population, except in the scenario where the cost associated with the control is low. In the latter scenario, we find strong differences across countries because the control extends to younger population in France and Vietnam 2-3 months after the onset of the epidemic, but not in Burkina Faso. Finally, we show that the optimal control strategy strongly outperforms constant uniform control of the whole population or over its younger fraction. This better understanding of the effect of age-based control interventions opens new perspectives for the field, especially for age-based contact tracing.

## 1 Introduction

Following its emergence in December 2019, COVID-19 has become an international public health emergency [1]. The infection has many similarities with that caused by influenza virus regarding clinical manifestations and transmission mechanism [1]. Contrary to seasonal influenza, COVID-19 has become pandemic by spreading rapidly among completely naive host populations, *i.e*. with no pre-existing immunity [2–5]. At the start of the pandemic, no pharmaceutical interventions such as vaccines or treatments were available and, based on earlier epidemics, it will take several months before their deployment. For this reason, non-pharmaceutical intervention strategies, such as social distancing, are key to controling the pandemic [6].

When an intervention is summarised by one or few parameter values, identifying an optimal strategy according to some criterion variable can readily be done, *e.g*. using a gradient approach [7]. Things become much more challenging when the intervention parameter value is a function of time. Optimal control theory [8], also known as Pontryagin’s maximum principle, specifically addresses this issue by identifying a function of time such that, over a finite time interval, some criterion is optimised. This has allowed studies to identify optimal non-pharmaceutical interventions to control infectious diseases such as influenza and COVID-19 [9–12]. However, a strong limitation of these studies is that they all ignore at least one aspect of the host population structure. First, infection parameters vary with infection age, *i.e*. depending on the number of days since infection. Second, hosts vary in age. The latter point is particularly important because in addition to being a function of time since the onset of the outbreak, optimal strategies involving physical distancing can also vary depending on host age [13–17]. Accounting for two dimensions, time and host age, make the optimisation procedure more challenging because Pontryagin’s maximum principle is applied to ordinary differential equations (ODEs) –something very common– while here we are working on partial differential equations (PDEs) –which is less common, and more challenging. Here, we address this challenge and identify interventions varying in intensity with time and host age, that significantly reduce morbidity associated with COVID-19 at a minimal cost. Furthermore, we compare the situation in countries with contrasted age-structure, namely Burkina-Faso, France, and Vietnam, to show how this affects optimal strategies.

The age structure of the population is a known key determinant of acute respiratory diseases, especially when it comes to infection severity. For example, children are considered to be responsible for most of the transmission of influenza virus [18], but the related hospitalization and mortality burden is largely carried by people of ages over 65 years [19, 20]. While much remains unknown about the COVID-19 epidemics, evidence to date suggests that mortality among people who have been tested positive for the coronavirus is substantially higher at older ages and near zero for young children [3, 21]. Moreover, the infectiousness of an individual has been reported to vary as a function of time since infection [22], which is known to affect epidemic spread [23–26].

Our epidemiological model for the disease stage-progression [24] is structured both by a (continuous) age of the host and a (continuous) age of infection. A variety of epidemiological models allow for one or the other type of structure [27–30], starting with a seminal article from the 1920s [23]. However, models allowing for a double continuous structure are rare [30–37]. Such a double structure is particularly suited to investigate an infection such as COVID-19, with strong host and infection age effects. Indeed, in addition to taking into account the age-structure of the host population, as well as the gradient of disease severity from mild to critical symptoms, the model readily captures the variation in infectiousness as a function of the time since infection. From a theoretical point of view, age-structured models have been proposed to investigate the spread of acute respiratory diseases [38–42], and some rare models of acute respiratory diseases consider both structures as continuous variables [30, 32], although not in the context of optimal control theory.

In Section 2, we first introduce the mathematical model. The model parameters and outputs are then defined in Section 3. In Section 4, we characterize the optimal control strategy that minimizes the number of deaths as well as the cost due to the implementation of the control strategy itself. Section 5 contains the main body of the results. We first analyse the epidemic spread without any intervention for three countries with contrasted age distributions (Burkina-Faso, France, and Vietnam). Next, the performance of optimal control in terms of deaths and hospitalizations is compared for different costs of the control measure. Finally, the optimal control is compared to two other strategies using the same amount of resources to control the outbreak. The article ends by a discussion in Section 6.

## 2 An age-structured epidemiological model

### 2.1 Model overview

We denote the density of individuals of age *a* ∈ [0, *a*_*max*_] that are susceptible to the infection at time *t* ∈ [0, *T*] by *S*(*t, a*). These individuals can become infected with a rate called the force of infection and denoted *λ* (*t, a*). We assume that a fraction *p* of these individuals are paucisymptomatic, which means that they will develop very mild to no symptoms, and enter group *I*_*p*_. Note that this class can also be interpreted as the fraction of the population that will not isolate themselves during their infection. Other individuals are assumed to develop more symptomatic infections, either severe *I*_*s*_ with proportion *q*(*a*) depending on the age *a*, or mild *I*_*m*_ with proportion 1 − *q*(*a*).

Each of the three infected host populations are structured in time since infection, so that *I*_*v*_(*t, a, i*), *v* ∈ {*p, s, m*}, denotes the density at time *t* of individuals of age *a* that have been infected for a duration *i* ∈ ℝ_+_. Upon infection, all exposed individuals are assumed to remain non-infectious during an average period *i*_*lat*_. Next, they enter an asymptomatic period during which they are infectious. Only *I*_*m*_ and *I*_*s*_ develop significant symptoms after an average time since infection *i*_*sympt*_, which can allow them to self-isolate to limit transmission. During their infection, individuals can recover at a rate *h*_*v*_(*a, i*) (*v* ∈ {*p, m, s*}) that depends on the severity of the infection and the time since infection *i*. Severely infected individuals of age *a* may also die from the infection at rate *γ*(*a, i*).

The infection life cycle is shown in Figure 1. The total size of the host population of age *a* at time *t* is

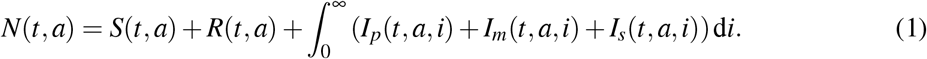

**Figure 1:**
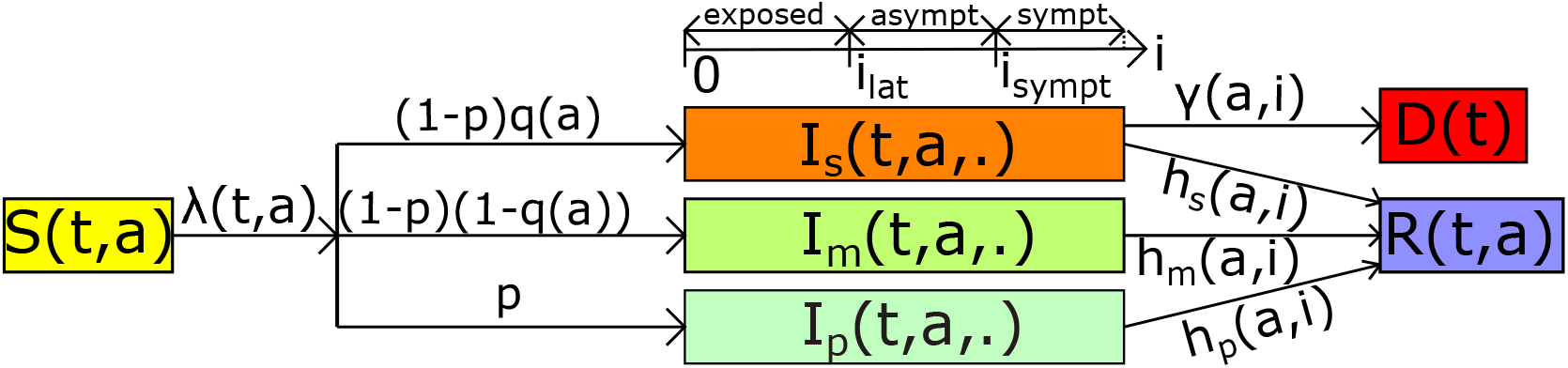
The model flow diagram. Susceptible hosts of age *a* at time *t* (*S*(*t, a*)) are exposed to the virus with a force of infection *λ* (*t, a*). A fraction *p* of exposed individuals, which are infected since time *i*, will never develop symptoms and enter the group of paucisymptomatic infections (*I*_*p*_(*t, a, i*)). The rest will develop symptomatic infections, either severe (*I*_*s*_(*t, a, i*)) with proportion *q*(*a*) depending on age *a* of individuals, or mild (*I*_*m*_(*t, a, i*)). Exposed individuals remain non-infectious for a duration *i*_*lat*_ after infection. Next, they become asymptomatic infectious and only symptomatic infected will develop symptoms at time *i*_*sympt*_ after infection. Infected individuals recover at rate *h*_*v*_(*a, i*). Only severely infected of age *a* die from the infection at rate *γ*(*a, i*). Notations are shown in Table 1.

#### Remark 2.1

*Contrarily to classical SEAIR models, disease-stage progression in our model is not captured by discrete compartments (exposed, asymptomatic, and infected) with exponentially distributed waiting times to switch between compartments. The advantage of our formalism is that disease progression can be modelled using a continuous variable, called the time since infection (in days) denoted here by i. Every infected person then remains in the “infection compartment” from exposure until recovery (or death). The time since infection grows linearly with time, according to the derivative with respect to i. Latency from exposed to asymptomatic and time of symptoms onset are not needed for this modelling approach because these are captured through the functions describing the transmission rate, the mortality rate, and the recovery rate at time i post infection. More precisely, the average latency from exposed to asymptomatic (i*_*lat*_*) is simply mentioned to define the average time to infection onset (i*_*sympt*_*), and also to help the readers to understand the model flow diagram (Figure 1). On the other hand, the time to infection onset (i*_*sympt*_*) is used to define infectiousness reduction factors (ξ* _*s*_, *ξ* _*m*_*) and the mortality rate due to the infection (γ)*.

### 2.2 Age-structured transmission and severity

We use two components to model the infection process. First, we define the transmission probability *β*_*v*_(*a, i*) (*v* ∈ {*p, m, s*}) for each contact between an infected of age *a* and a susceptible person, which depends on the time since infection *i*. Second, we introduce the kernel *K*(*a, a*^′^) that represents the average number of contacts by unit of time between an individual of age *a*^′^ and an individual of age *a*.

Here, this contact matrix is informed by data from earlier study [43]. The force of infection underwent by susceptible individuals of age *a* at time *t* is then given by

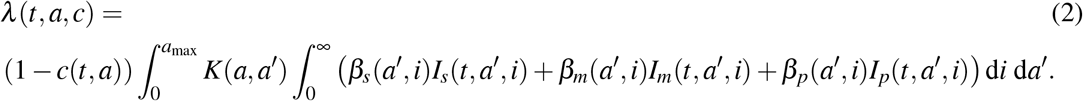

Here, *c* = *c*(*t, a*) is the percentage of contacts reduction towards people with age *a*, due to public measures, at time *t*. The total force of infection at time *t* in the whole population is computed as 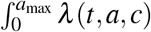d*a*. The dynamics of newly infected individuals (*i.e. i* = 0) in each group is thus defined by

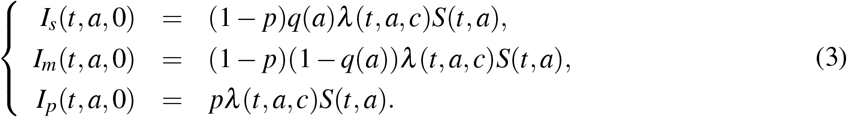

Note that *p* likely depends on age, but because is it totally unknown, we assume it is a constant. Further, we assume that only severe infections *I*_*s*_ lead to hospitalization and we denote by

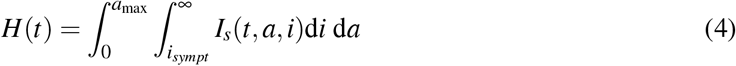

the total population hospitalized at time *t*, where *i*_*sympt*_ is the average time to symptoms onset. Each individual of age *a* dies at a rate *µ*(*a, H*(*t*)) at time *t*, defined by

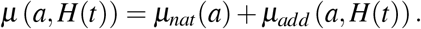

In the latter equation, *µ*_*nat*_ denotes the natural mortality rate when hospitals are not saturated. Further, we assume that this rate increases significantly as soon as the number of severe cases exceeds the healthcare capacity *H*_*sat*_ and *µ*_*add*_ is such additional death rate due to hospital saturation (see Section 3.2).

We apply the same reasoning by assuming that the disease-related mortality can increase because of hospital saturation. Therefore, severely infected individuals of age *a* infected since time *i* die at time *t* at rate *γ*(*a, i, H*(*t*)) defined by

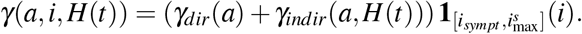

Here, *γ*_*dir*_ and *γ*_*indir*_ are mortality rates directly and indirectly due to the COVID-19 respectively (see Section 3.2). The disease-related mortality occurs after the emergence of symptoms and before the mean final time of infection for severe cases, *i.e*. for 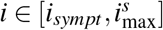.

Finally, infected individuals of age *a* infected since time *i* recover at rates *h*_*s*_(*a, i*), *h*_*m*_(*a, i*) and *h*_*p*_(*a, i*) for severe, mild and paucisymptomatic infections respectively.

The boundary conditions (3) are coupled with the following equations:

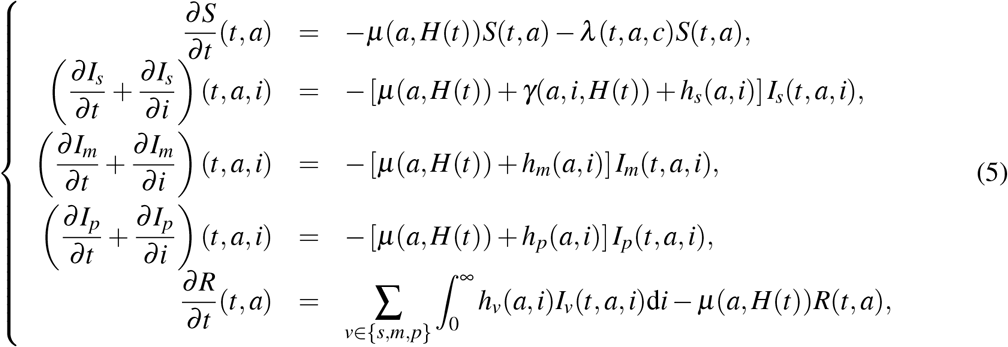

for any (*t, a, i*) ∈ (0, *T*] × [0, *a*_max_] × ℝ_+_, with initial conditions (at *t* = 0):

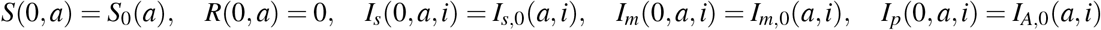

for each (*a,i*) ∈ [0, *a*_max_] × ℝ_+_. The initial conditions of infected populations are detailed in Section 3.3. Using (3) and an integration over *i* of (5), one may observe that the total population *N* defined by (1) is strictly decreasing since it satisfies the following inequality:

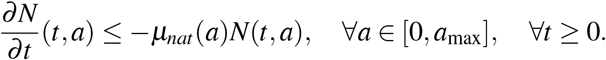

This is due to the fact that population aging and births are neglected in this model since we consider a time horizon of only one year. Further, basic properties of the model such as existence and positiveness of solutions is out of the primary scope of our study. However, these can be specifically addressed using an integrated semigroup approach and Volterra integral formulation (see *e.g*. [44–47] and references therein). More specifically, one may follow [31] where the well-posedness of an epidemiological model with a double continuous structure is handled.

## 3 Epidemiological outputs, model parameters and initial conditions

In this section we briefly describe some useful epidemiological outputs, the shape of age-dependent parameters considered for the simulations of model (3)-(5), and the initial conditions. All state variables and other parameters are summarized in Table 1.

**Table 1:** Model variables and parameters.

| Param. | Description (unit) | Values [source] |  |  |
| --- | --- | --- | --- | --- |
| State variables |  |  |  |  |
| $S$ | Susceptible individuals | | | |
| $I_s$ | Severely infected individuals | | | |
| $I_m$ | Mildly infected individuals | | | |
| $I_p$ | Paucisymptomatic infected individuals | | | |
| $R$ | Recovered individuals | | | |
| General parameters |  |  |  |  |
| $t, T$ | time and final time of simulations (days) | $t \in [0, T]$ (ad hoc) | | |
| $a, a_{\max}$ | age and maximal age of individuals (years) | $a \in [0, a_{\max}]$, $a_{\max} = 100$ (ad hoc) | | |
| $i$ | time since infection (days) | $\mathbb{R}_+$ (ad hoc) | | |
| $i_{lat}$ | average latency from exposed to asympt. (days) | 4.2 [49] | | |
| $i_{symp}$ | average time of symptoms onset (days) | $i_{lat} + 1 = 5.2$ [48] | | |
| $i_{\max}^s$ | mean final time of infection for severe cases (days) | $i_{symp} + 20 = 25.2$ [50] | | |
| $i_{\max}^m$ | mean final time of infection for mild cases (days) | $i_{symp} + 17 = 22.2$ [50] | | |
| $\mu_{add}$ | additional death rate ($\text{days}^{-1}$) | defined by (8) | | |
| $\beta_s, \beta_m, \beta_p$ | transmission probabilities (unitless) | computed in Section 3.2 | | |
| $\xi_s, \xi_m, \xi_p$ | infectiousness reduction factors (unitless) | defined by (9) and $\xi_p = 0.1$ [22] | | |
| $h_s, h_m, h_p$ | recovery rates per infection ($\text{days}^{-1}$) | defined by (10) | | |
| $c, c_{\max}$ | percentage of contacts reduction and its upper bound | $c \in [0, c_{\max}]$, $c_{\max} = 0.95$ (assumed) | | |
| $\gamma_{dir}$ | mortality rate directly due to the COVID-19 ($\text{days}^{-1}$) | [48] | | |
| $\gamma_{indir}$ | mortality rate indirectly due to the COVID-19 ($\text{days}^{-1}$) | defined by (8) | | |
| $p$ | proportion of paucisymptomatic (unitless) | variable | | |
| $q$ | proportion of symptomatic requiring hospitalisation (unitless) | [48] | | |
| $B$ | cost of the control measure (unitless) | variable | | |
| Specific parameters for each country |  |  |  |  |
| Param. | Description (unit) | Burkina Faso | France | Vietnam |
| $S_0$ | initial population of susceptible | [51] | [52, 53] | [54] |
| $I_0^{(*)}$ | initial epidemic size | 288 (WHO)(**) | 130 [55] | 217 (Ministry of Health) |
| $\mu_{nat}$ | natural death rate ($\text{days}^{-1}$) | [56] | [57] | [58] |
| $H_{sat}$ | maximal healthcare capacity (unitless) | 11 [59] | 5000 [55] | 5932 (NIHE)(**) |
| $K$ | matrix of social contacts ($\text{days}^{-1}$) | [43] | [43] | [43] |
| Parameters and range for the global sensitivity analysis |  |  |  |  |
| Param. | Description | Range |  |  |
| Pop.Struc | population structure | {Burkina Faso, France, Vietnam} |  |  |
| $H_{sat}$ | maximal healthcare capacity | {10, 100, 500, 2000, 5000, 6000, 50000, 5e+05, 5e+06} | | |
| $p$ | proportion of paucisymptomatic | {0.05 to 0.95} by step of 0.1 | | |
| $i_{symp}$ | average time of symptoms onset | {1.2 to 9.2} by step of 2 | | |
| $\xi_p$ | infectiousness reduction of $I_p$ | {0.1, 0.3, 0.5, 0.7, 1} | | |

(\*) : corresponds to March, 1st, 2020 in France and April, 1st, 2020 in Burkina Faso and Vietnam.
(\*\*) : WHO: World Health Organisation, NIHE: National Institut of Hygiene and Epidemiology

### 3.1 Epidemiological outputs

In addition to the total number of hospitalized cases *H*(*t*) at time *t* defined by (4), we define additional epidemiological outputs such as the number of non-hospitalized cases (*N*_*H*_(*t*))

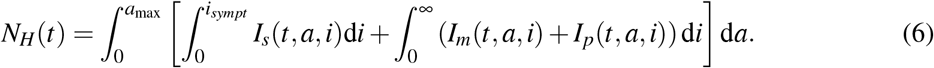

Note that the latter encompasses paucisymptomatic, mildly infected, and severely infected but not yet hospitalized hosts.

For the cumulative number of deaths, we distinguish between those directly due to COVID-19 Infections 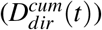, and those indirectly due to the epidemic 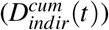, which originate from the saturation of the health system:

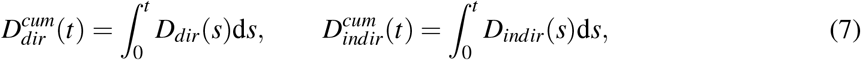

where *D*_*dir*_(*t*) and *D*_*indir*_(*t*) are the number of deaths at time *t* respectively defined by

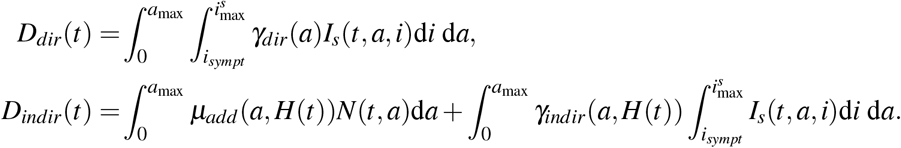

Every aforementioned output implicitly depends on parameter *c* = *c*(*t, a*), which we will omit in the notations when no confusion is possible. However, for clarity, we do explicitly write this dependence to compare public health measures. The relative performance between two strategies *c*_1_ and *c*_2_, denoted by Δ(*c*_1_, *c*_2_), is estimated by assessing the cumulative number of deaths in the whole population during the *T* days of control period with the strategy *c*_1_ relatively to deaths with the strategy *c*_2_. Formally we have

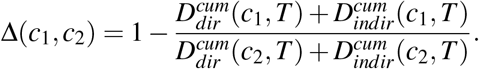

Hence, a relative performance Δ(*c*_1_, *c*_2_) = 0.1 implies that the strategy *c*_1_ reduces the number of deaths by 10% relatively to *c*_2_.

### 3.2 Model parameters

#### Mortality rates

We assume that indirect mortality, *i.e*. not directly due to COVID-19, increases when the number of hospitalisations *H*(*t*), at time *t*, exceeds a healthcare capacity threshold *H*_*sat*_ (which is approximated with the maximal intensive care capacity). The natural mortality rate then increases by *µ*_*add*_(*a, H*) for the whole population, and by *γ*_*indir*_(*a, H*) for severely infected individuals of age *a*. These rates are modelled by logistic functions that are arbitrarily chosen as:

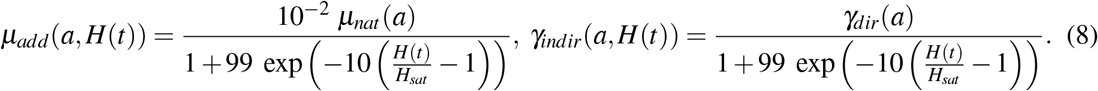

This choice of functional parameters implies that

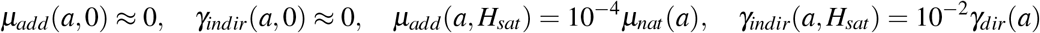

which means these additional mortalities are negligible when hospitals are not saturated (Figure 2 b). In case of full saturation, we have

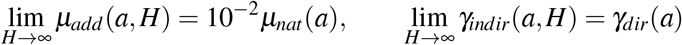

for each *a* ∈ [0, *a*_max_], meaning that the natural mortality rate is only increased by 1%, while the disease-induced mortality rate *γ* is doubled. Indeed, according to [48], less than 50% of patients in critical care will die in case of no saturation of hospitals.

**Figure 2:**
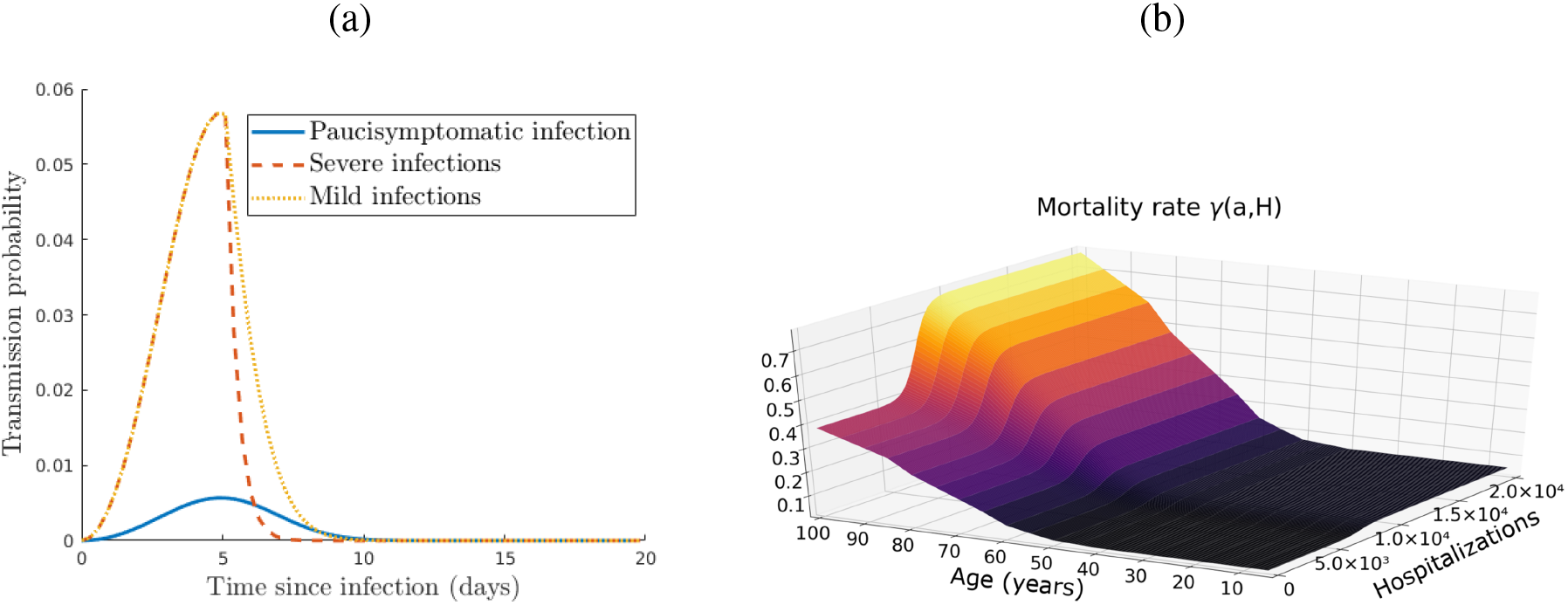
(a) Transmission probabilities of paucisymptomatic infections *β*_*p*_, symptomatic severe *β*_*s*_ and mild infections *β*_*m*_. (b) Disease induced mortality rate with a maximal healthcare capacity *H*_*sat*_ = 5 × 10^3^.

#### Transmission rates

The infectiousness of an individual aged *a*, which is infected since time *i*, is given by *β*_*v*_(*a, i*) (*v* ∈ {*s, m, p*}). Based on estimates described in [22], we assume that *β*_*v*_ does not depend on host age *a, i.e*., *β*_*v*_(*a, i*) = *β*_*v*_(*i*). This assumption is only made for parameterization purpose and does not impact the general formulation of the model proposed here (this is discussed later in Section 6).

The transmission rate at a given day *i* post infection of a given type of infectious host is defined such that 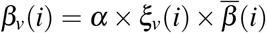, for *v* ∈ {*s, m, p*}. As detailed below, *α* is a scaling parameter obtained from the value of the basic reproduction number *R*_0_, which is the mean number of secondary infections caused by an infected host [24]. As [22], we assume that parameter 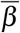, which strongly depends on the generation interval, follows a Weibull distribution 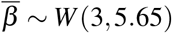. Finally, parameters *ξ* _*v*_(*i*) are factors that capture variations in infectiousness based on the type of host. For paucisymptomatic individuals, for instance, these are assumed to be constant (*ξ* _*p*_(*i*) = *ξ* _*p*_), while the reduction factor in more symptomatic infections (severe and mild) is assumed to vary after symptom onset to capture admission in a healthcare facility or self-isolation at home. More precisely, we assume that

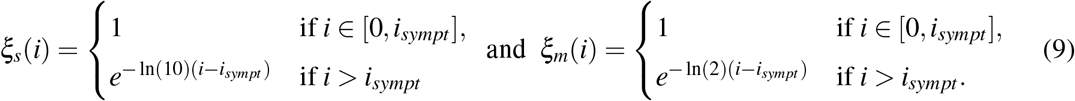

These two functions are chosen arbitrarily by assuming that individuals do not isolate before symptoms onset (*i* ≤ *i*_*sympt*_), and that isolation is stronger when symptoms are more severe (Figure 2 a). We therefore assume that the transmission probability 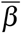 is divided by 10 (respectively 2) every day after the average time of symptoms onset for individuals severely (resp. mildly) infected.

#### Recovery rates

We assume that recovery rates *h*_*v*_(*a, i*), *v* ∈ {*s, m, p*}, of infected individuals of age *a* infected since time *i* are independent of the age *a* and take the following form:

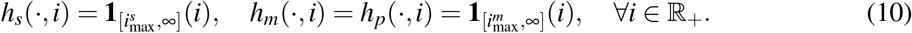

That is, one can recover from severe (resp. mild and paucisymptomatic) infections only after a time since infection 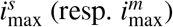 corresponding to the mean duration of infection.

### 3.3 Initial conditions

The initial susceptible population *S*_0_ and epidemic size *I*_0_ are given in Table 1. Since, initially, screening is usually restricted to individuals with severe symptoms, we assume that all initial cases are severe infections. Thus, we set 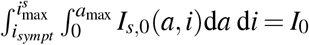 as the initial severely infected individuals, which we assume to be uniformly distributed with respect to the time since infection *i* on the interval 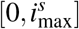. Using estimates from [55, 60] on the age distribution of hospitalised people, we derive an estimation of *I*_*s*,0_(*a, i*) for each (*a, i*) ∈ [0, *a*_max_] × ℝ_+_. Next, following the life cycle (Figure 1), we obtain an estimation of the total initial infected population by 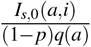. From there, we deduce the initial mildly and paucisymptomatic infected populations, which can be denoted respectively by

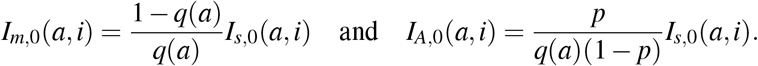

## 4 Optimal intervention

As explained above, our goal is to find an optimal control strategy that is allowed to vary depending on the number of days since the onset of the epidemic (*t*) and on host age (*a*). In this section, following well established methodology in optimal control theory [13–16, 61], we search for the optimal control effort function *c*\* that minimizes the objective functional *J* : *L*^∞^(ℝ_+_ × [0, *a*_max_]) ∋ *c* ↦ *J*(*c*) ∈ ℝ, where

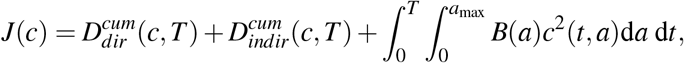

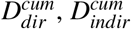 being the cumulative number of deaths defined by (7), and *B*(*a*) the cost associated with the implementation of such control *c* for the age class *a*. Our aim is to find the function *c*\* satisfying

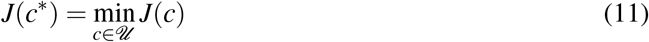

wherein the set *𝓊* is defined by

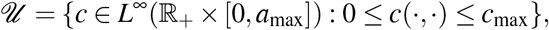

with *c*_max_ ≤ 1 a positive constant. That is to say, the function *c*\* will minimize the cumulative number of deaths during *T* days, as long as the cost of the control strategy is not too large.

Let (*S, I*_*s*_, *I*_*m*_, *I*_*p*_, *R*) be a given solution of (3)-(5) then let *λ* and *H* be respectively defined by (2) and (4). After some computations (Appendix C), we find that the adjoint system of (5) reads as

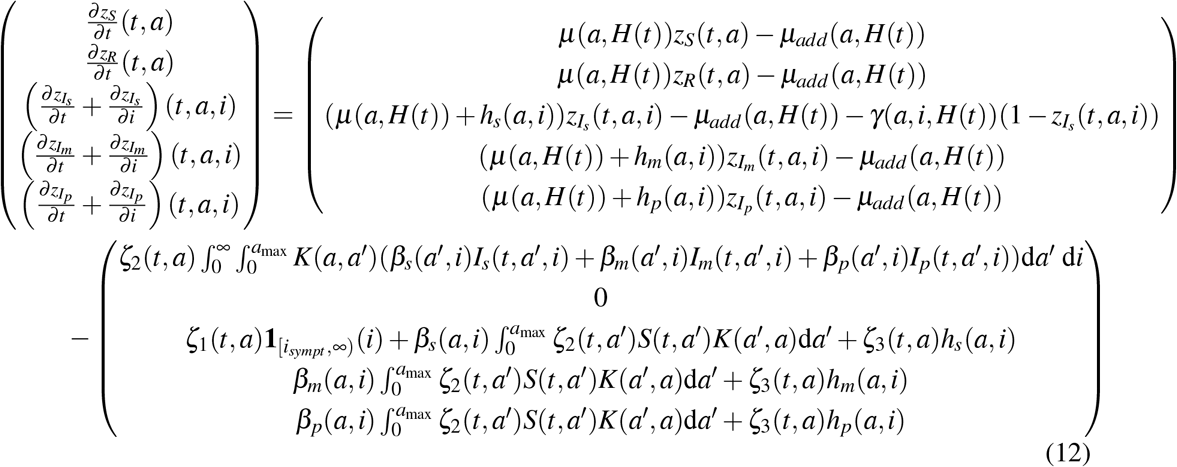

with final conditions *z*_*S*_(*T, a*) = *z*_*R*_(*T, a*) = 0, *z*_*u*_(*T, a, i*) = 0 and lim_*i*→∞_ *z*_*u*_(*t, a, i*) = 0, for any *u* ∈ {*I*_*s*_, *I*_*m*_, *I*_*p*_} and (*a, i*) ∈ [0, *a*_max_] × ℝ_+_, while *ζ* _*k*_ (*k* ∈ {1, 2, 3}) satisfy the system:

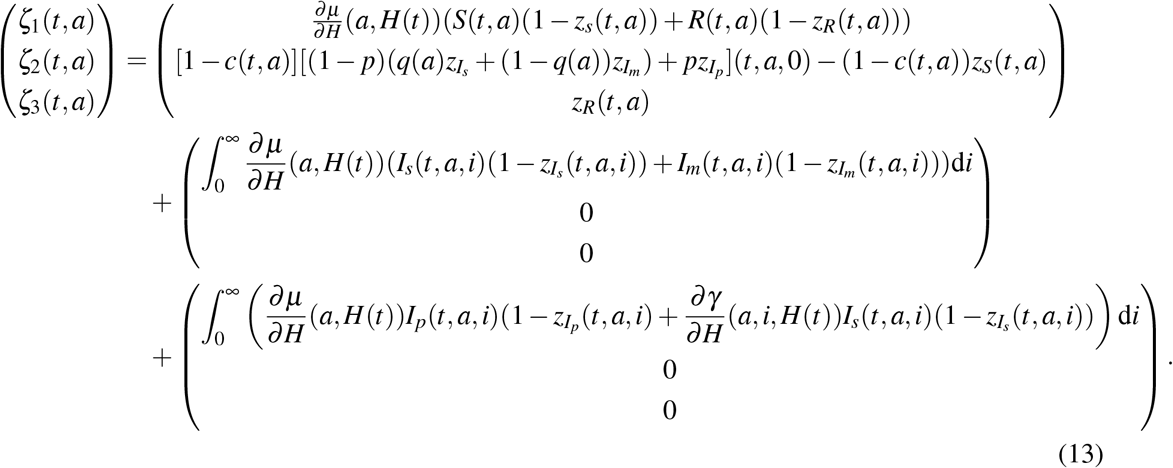

Finally, the Hamiltonian *ℋ* of (11) is given by (C.1). Then, solving 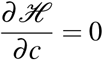, it comes that

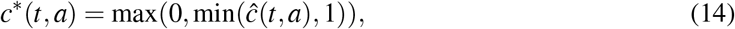

for every (*t,a*) ∈ [0;*T*] × [0;*a*_max_], where

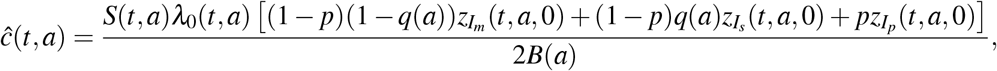

with *λ*_0_ defined by (B.4).

We also assume that the cost *B*(*a*) of the control measure over individuals aged *a* ∈ [0, *a*_max_] is proportional to their density in the initial susceptible population *S*_0_, *i.e*.

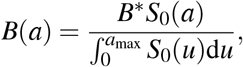

where *B*\* ∈ ℝ_+_ is a variable parameter characterizing the relative cost in implementing the strategy. Additionally, one may consider the age distribution of the economic cost on the shape of the function *B*. For example, the economic cost can be assumed more important for the working population (*i.e*. age group 20 − 60) compared to the older, mostly retired, population. However, in absence of relevant references on this topic we stand with our primary assumption.

The state system (3)-(5) and the adjoint system (12)-(13) together with the control characterization (14) form the optimality system to be solved numerically. Since the state equations have initial conditions and the adjoint equations have final time conditions, we cannot solve the optimality system directly by only sweeping forward in time. Thus, an iterative algorithm, forward-backward sweep method, is used [8]. In other words, finding *c*\* numerically, involves first solving the state variables (3)-(5) forward in time, then solving the adjoint variables (12)-(13) backward in time, and then plugging the solutions for the relevant state and adjoint variables into (14), subject to bounds on the control function. Finally, the proof of the existence of such control is standard and is mostly based on the Ekeland’s variational principle [62]. Therefore, existence of the optimal control to the above problem is assumed and we refer to [13] for more details.

## 5 Results

Here we consider three countries as case studies: Burkina Faso, France and Vietnam. They have quite contrasted age-structure and social contacts of their population (Figure 3). Indeed, in Burkina Faso the very large majority (96.1 %) of the population is under 60 while it is respectively 87.7 % and 73.4 % in Vietnam and France (Figure 3 a,b). It shows that an higher proportion of the population is older than 60, hence at risk for COVID-19 infection, in France 26.6%, in comparison to Vietnam 12.3 %, or to Burkina Faso 3.9 % (Figure 3 a,b). Also, contacts are more frequent among the older population in France compared to Vietnam (Figure 3 d, e). By contrast, very few contacts are observed among older populations in Burkina Faso (Figure 3 c).

**Figure 3:**
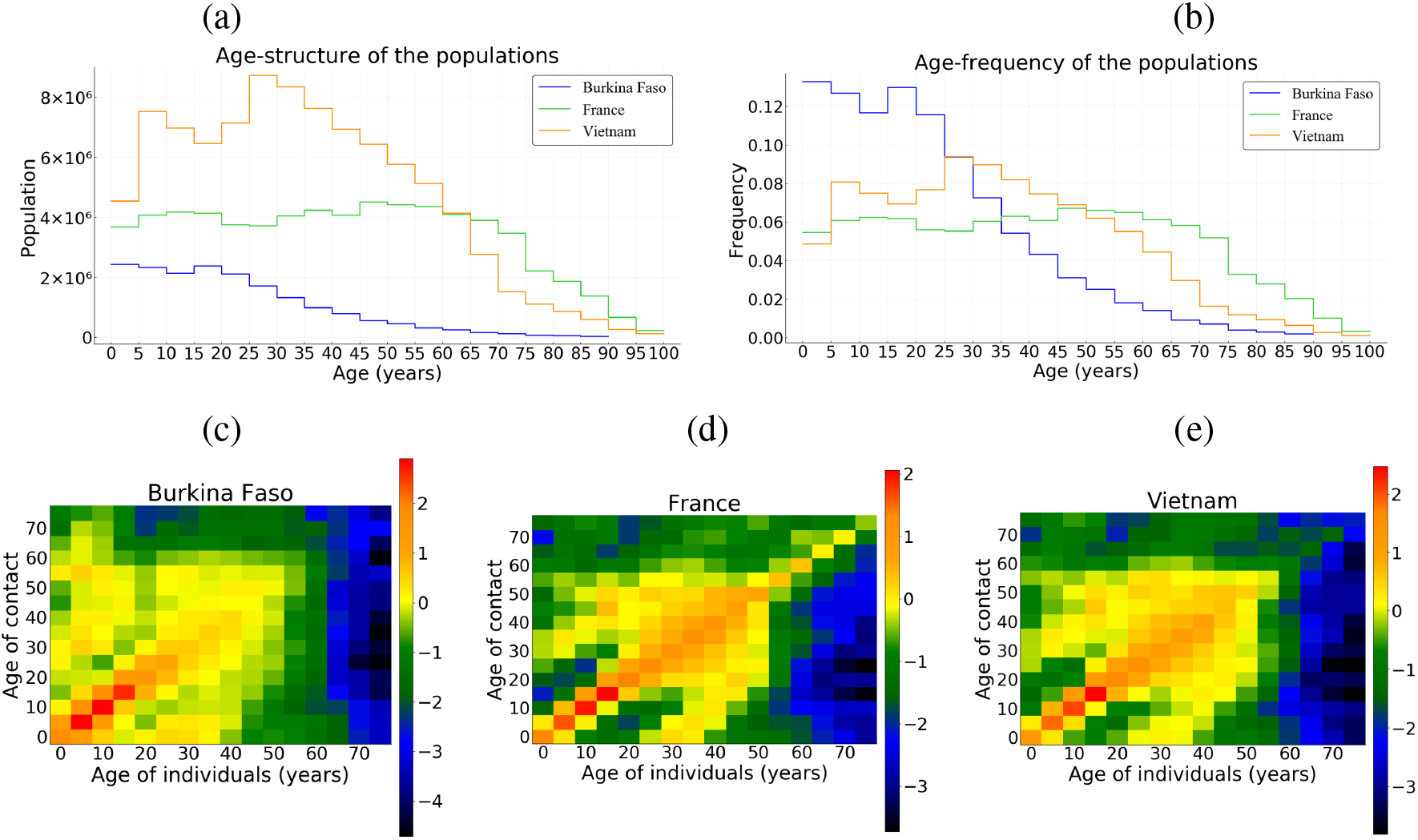
(a)-(b) The population age-structure of Burkina Faso, France and Vietnam. (c)-(e) Contact matrices in the three countries, in log scale where dark color intensities indicate less likely events *i.e*. smaller tendency of having a household member of that age, lower proclivity of making the agespecific contact.

### 5.1 Global sensitivity analysis

We study the sensitivity of infected individuals, hospitalizations and deaths to five parameters: proportion of paucisymptomatic (*p*), average time of symptoms onset (*i*_*sympt*_), infectiousness reduction of paucisymptomatic infections (*ξ* _*p*_), healthcare capacity (*H*_*sat*_) and population structure (including the natural mortality, the size of the population, age-structure and social contacts). The variation range of above parameters is assigned in Table 1. Sensitivity indices are estimated by fitting an ANOVA (Analysis Of Variance) linear model, including third-order interactions, to the data generated by simulation. Note that this ANOVA linear model fitted well with 99% of variance explained. Overall, the population structure is the main parameter highlighted by the sensitivity analysis with 70% of the variance explained for the number infected individuals, 40% for hospitalizations and deaths (Figure S1). The population structure is followed by *ξ* _*p*_, *p*, and *i*_*sympt*_ which have quite similar importance on the number infected individuals with a slight dominance of *ξ* _*p*_ (Figure S1). By contrast, for hospitalizations and deaths, the population structure is followed by *p* with 40% and 30% of the variance explained respectively; while *ξ* _*p*_ and *i*_*sympt*_ have very marginal impact (Figure S1). Finally, the importance of *H*_*sat*_ is strictly negligible on the three output variables, with however, a greater importance on deaths as compared to hospitalizations and infected (Figure S1).

### 5.2 The basic reproduction number *R*_0_

An explicit expression of the *R*_0_ of model (3)-(5) is difficult to obtain in general. We show in Appendix B that it is possible to write *R*_0_ = α × *r* (*Ū*), where *α* is the scaling parameter introduced in Section 3.2, and *r(Ū*) is the spectral radius of the next generation operator *Ū* defined on *L*^1^(0, *a*_max_) by

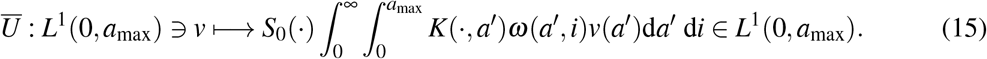

where *S*_0_ is the initial susceptible population, *K* is the contact matrix and *ω*(*a, i*) is the infectiousness of individuals of age *a* infected since time *i* (Appendix B). It follows that

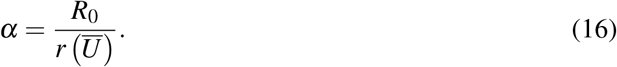

Setting *R*_0_ = 3.3 [63, 64] for all three countries and using a numerical approach and corresponding values for *S*_0_ and *K* for each country, we successively determine *r*(*Ū*) and *α* by (15) and (16) respectively.

### 5.3 Uncontrolled epidemic

We first use the model (3)-(5) to describe the outbreak of the epidemics for all three countries, without any public health measure (*i.e. c* ≡ 0), with *R*_0_ = 3.3 and other parameters defined in Section 3 and summarized in Table 1.

The peak of the epidemics is reached approximately at day *t* = 51 for hospitalised people, and day *t* = 46 for non-hospitalised people without any control measures in the France scenario (Figure 4 e). Such times to peaks for hospitalised and non-hospitalised people are 47 and 41 (resp. 50 and 45) for Burkina Faso (resp. Vietnam) scenario (Figure 4 a, resp. Figure 4 i). The delay between the two peaks is due to the latency time *i*_*sympt*_ for symptoms onset (Table 1).

**Figure 4:**
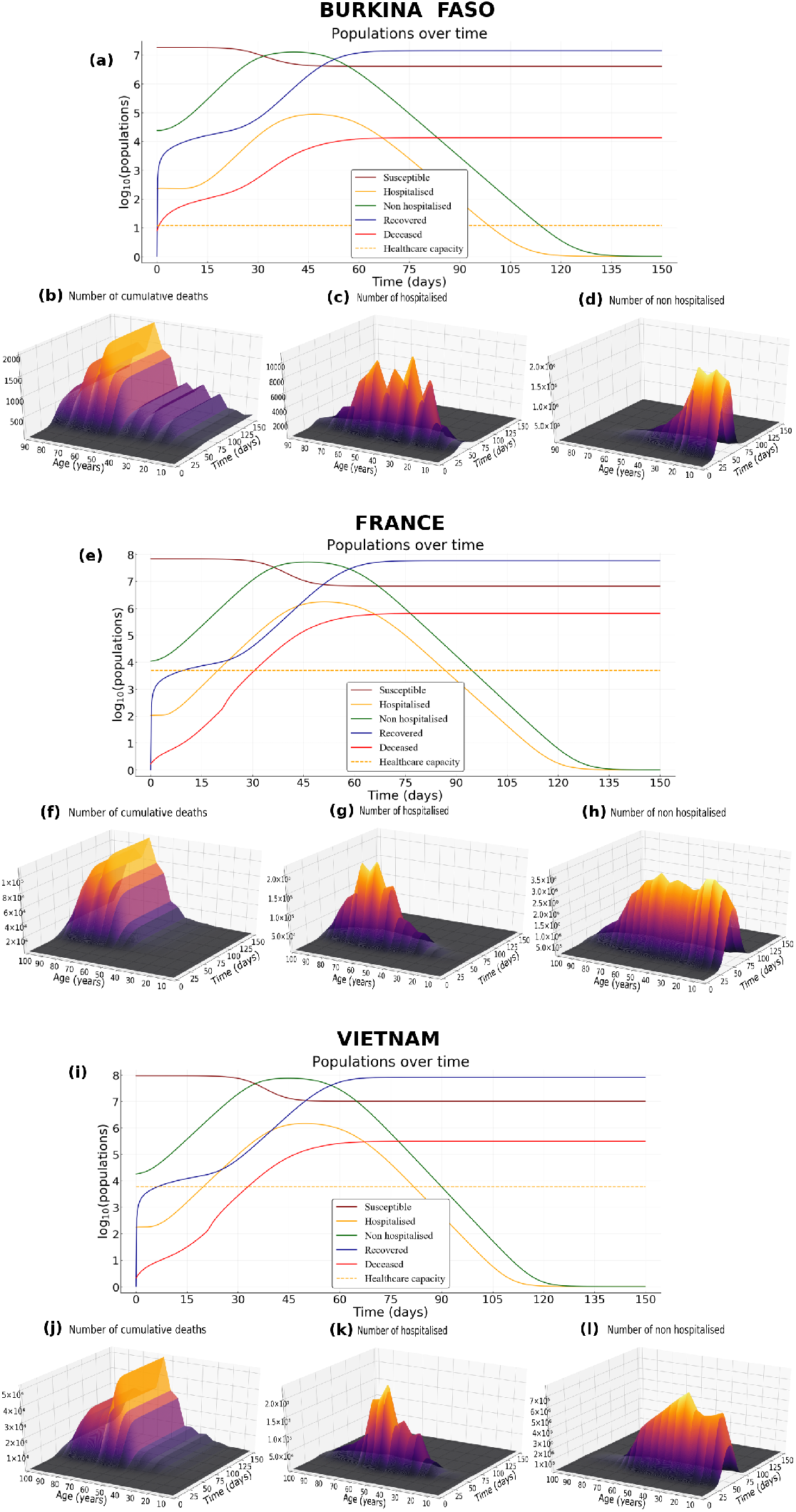
Epidemic scenario without any control measures. (a) Dynamics of epidemiological outputs, (b) number of cumulative deaths, (c) number of hospitalised and (d) non-hospitalised people in Burkina Faso. (e-h) As for (a-d) but in France. (i-l) As for (a-d) but in Vietnam. Parameter values are default in Table 1, *R*_0_ = 3.3 and the proportion of paucisymptomatic infections is *p* = 0.5.

In absence of control measures, the healthcare capacity is quickly exceeded, about twenty days for France scenario (Figure 4 e), and the number of deaths increases sharply from then on. Such configuration is similar for Vietnam scenario (Figure 4 i). By contrast, due to a very low healthcare capacity in Burkina Faso, the health system is exceeded only after a few days compared to France and Vietnam (Figure 4 a). However, this overloading of the health system does not have the same consequences in terms of mortality in Burkina Faso compared to France and Vietnam. This is partially explained on the one hand by the fact that less than 4% of the population is above 60 years in Burkina Faso (Figure 3 a) and on the other hand by the fact that very few contacts are observed with older population in Burkina Faso compared to France or Vietnam (Figure 3 b-d).

At the end of the simulation (*t* = 150 days), without any control measures, the herd immunity threshold (1 − 1*/R*_0_ ≈ 69.7%) is reached in Burkina Faso, France and Vietnam (Figure 5). Indeed, the average size of the epidemic (severe, mild, and paucisymptomatic infections) is close to 90% in France and Vietnam but only 78% in Burkina Faso (Figure 5). Interestingly, in all three countries, the proportion of the population less than 20 that have been infected is around 93%. While almost the same proportion of the group [20 − 60] was infected in France and Vietnam (94%), only 65% was infected in Burkina Faso. This proportion then decreases for the population older than 60, more or less quickly depending on the country, and is around 73% in France, 56% in Vietnam and 33% in Burkina Faso. Further, among the infected population, more than 98% are less than 60 in Burkina Faso, while this proportion is 92% in Vietnam and 76% in France. This age structure of infected populations is particularly important since most of the infections that occur in the young population do not require hospitalisation (Figures 4g, 4 c, 4 k) while people older than 60 represent the age class with the highest cumulative number of deaths (Figures 4 f, 4 b, 4 j).

**Figure 5:**
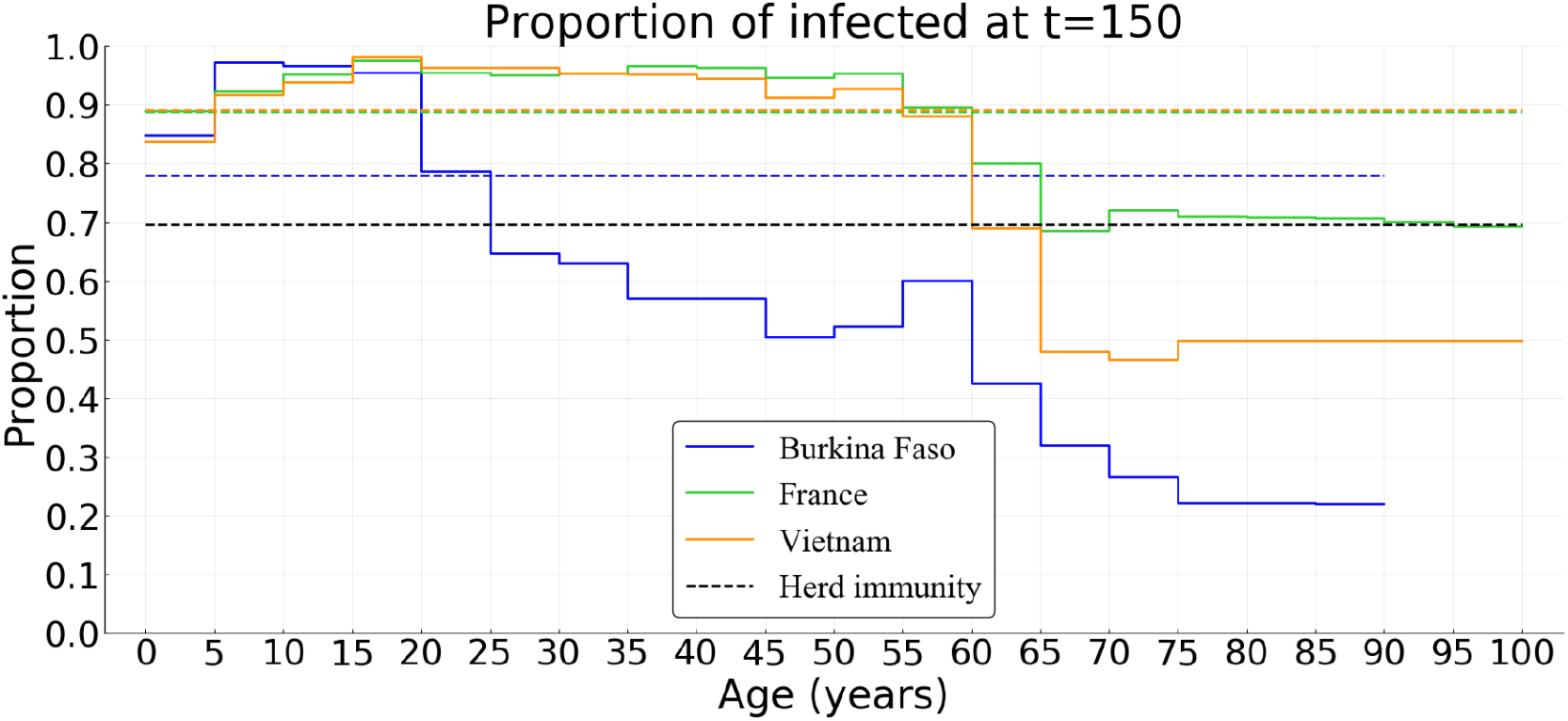
Simulated age distribution of the proportion of the population infected in Burkina Faso, France, and Vietnam in absence of control measures. Parameter values are default, *R*_0_ = 3.3 and the proportion of paucisymptomatic infections is *p* = 0.5.

### 5.4 Optimal intervention

We now investigate the result of implementing an optimal intervention that accounts for the age structure of the population. Strategies performances are here compared in terms of cumulative number of deaths for three costs of control measures (low *B*\* = 10^2^, intermediate *B*\* = 10^3^, and high *B*\* = 10^4^).

The optimal control strategy varies in time and depends on host age. In general, regardless of the country (Burkina Faso, France or Vietnam), the control is stronger early in the epidemic and for older populations (Figures 6, S2, S3). Overall, the level of optimal control is lower in Burkina Faso compared to France and Vietnam (Figures 6, S2, S3). If the cost of implementing the measures *B*\* is intermediate or high, the optimal control is almost restricted to individuals above 55 and to the first third of the time interval considered, with a significant reduction in deaths (Figures 6 d, e, Figure S2 d, e and Figure S3 d, e). In France, the relative performance of the optimal control *c*\* compared to a ‘doing nothing’ scenario (Δ(*c*\*, 0)) is at least 92% (resp. 82%) when the cost is *B*\* = 10^3^ (resp. 10^4^). For Burkina Faso, Δ(*c*\*, 0) is at least 50% (resp. 4%) when *B*\* = 10^3^ (resp. 10^4^). Finally, for Vietnam Δ(*c*\*, 0) is at least 87% (resp. 62%) when *B*\* = 10^3^ (resp. 10^4^). In the case of Burkina Faso, note that the level of the optimal control is quite low when the cost of implementation is high (Figure S2 c), and as a result, the effect of this control in reducing mortality at the population level is negligible. This is due to the relatively small number of deaths in the whole population in Burkina Faso without any control measures (Figure 4 a).

**Figure 6:**
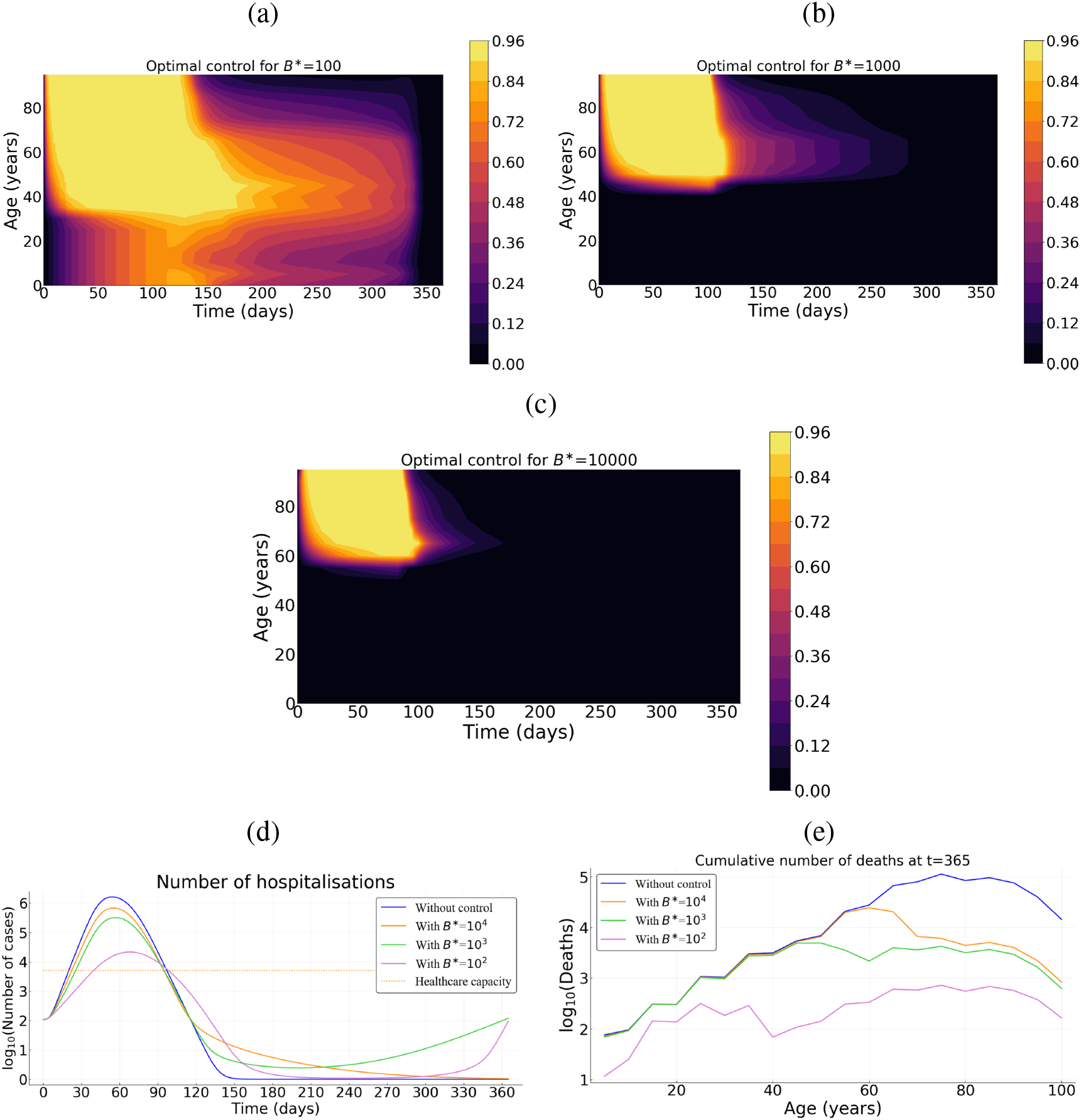
Optimal control strategy (*c*\*) as a function of the cost of the control measures in France. Intensity of the control as a function of time and host for for (a) relatively low *B*\* = 10^2^, (b) an intermediate *B*\* = 10^3^, and (c) a high *B*\* = 10^4^ cost. (d) Prevalence of hospitalized patients as a function of the strategy and the cost. (e) Cumulative deaths per age at the end of the time interval (when *T* = 365 days). Parameter values not related to the control are identical to Figure 4. Cases of Burkina Faso and Vietnam are given by Figures S2, S3.

If the implementation of the control measure comes at a low cost (*B*\* = 10^2^), the optimal control significantly extends to younger populations in all three countries (Figures 6 a, S2 a, S3 a), with a maximum intensity reached near the 4th month of the epidemics and a steady decrease until the end of the control period. Overall, the optimal control lasts less longer in Burkina Faso (Figure S2 a) compared to the cases of France and Vietnam (Figures 6 a, S3 a). At first, the control is mainly applied to people above 35 in all three countries (Figures 6 a, S2 a, S3 a). But, while the control extends to people less than 35 in France and Vietnam after 2 or 3 months (Figures 6 a, S3 a), such an extension is very moderate (or even negligible) in Burkina Faso (Figure S2 a). The resulting reduction in the number of deaths is very pronounced with a relative performance Δ(*c*\*, 0) of at least 80% (resp. 99%, 97%) in Burkina Faso (resp. France, Vietnam).

### 5.5 Performance and practical implementation

To illustrate how the strategy identified using optimal control theory outperforms “classical” optimisation approaches, we derive optimal strategies that do not vary in time and use the same amount of “resources” (that is the same cumulative cost). Assuming a relatively high cost *B*\* = 10^3^, we first investigate a control strategy that targets the younger fraction of the population (Figure 7 a), a second strategy that uniformly targets the whole population (Figure 7 b). Both strategies have a control level *c*_max_ = 0.95 and vary in duration (the total amount of resource used being constant).

**Figure 7:**
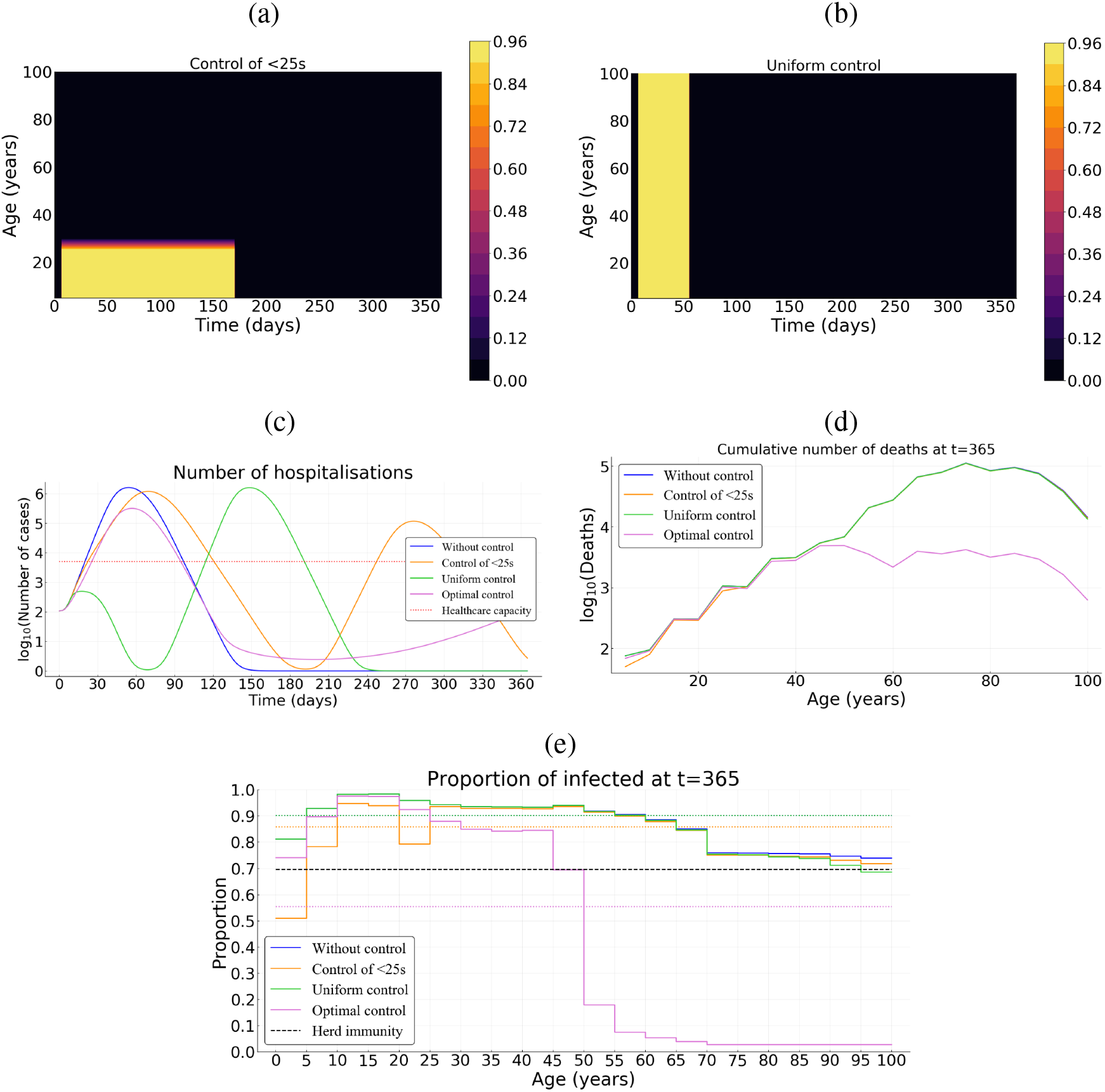
Comparing optimal control with uniform control of the whole population of over its younger fraction in France. (a) Illustration of the control over the young population and (b) uniform control of the whole population. (c) Number of hospitalizations. (d) Cumulative deaths per age at final time *T* = 365 days. (e) Age distribution of the proportions of the population that have been infected before one year. Here, we assume *B*\* = 10^3^ and *p* = 0.5.

In France, when targeting the population uniformly, the epidemic is under control during approximately 60 days. However, once the control resources are exhausted, the epidemic reemerges (Figure 7 c). With the (longer) control over the younger fraction of the population, the first epidemic peak is slightly delayed and the epidemic appears to be under control for a longer time period (180 days). Unfortunately, resources also become exhausted and a second peak appears a few months later (Figure 7 c). Whether it is for a uniform control of the whole population or over its younger fraction, the cumulative mortality over the time period of interest is comparable to that without any control measure (Figure 7 d). The performance of the optimal control relatively to the uniform control of the whole population or over its younger fraction, is approximately 92%; and at the end, 55% of the whole population has been infected with the optimal control and at least 85% with control of the whole population or over its younger fraction (Figure 7 e). A such configuration is quite similar for the case of Vietnam (Figure S4).

By contrast, for the case of Burkina Faso, regardless the control strategy (optimal, uniform or over the younger fraction) the proportion of infected population is approximately the same as without control (78%). The herd immunity threshold (1 − 1*/R*_0_ ≈ 69.7%) is then reached for all the three control measures and the epidemic cannot restart (Figure 8 e). The cumulative mortality with a uniform control of the whole population or over its younger fraction is comparable to that without any control measure (Figure 8 d). However, despite their same proportion of infected individuals, the performance of the optimal control relatively to the uniform control of the whole population or over its younger fraction, is at least 50%.

**Figure 8:**
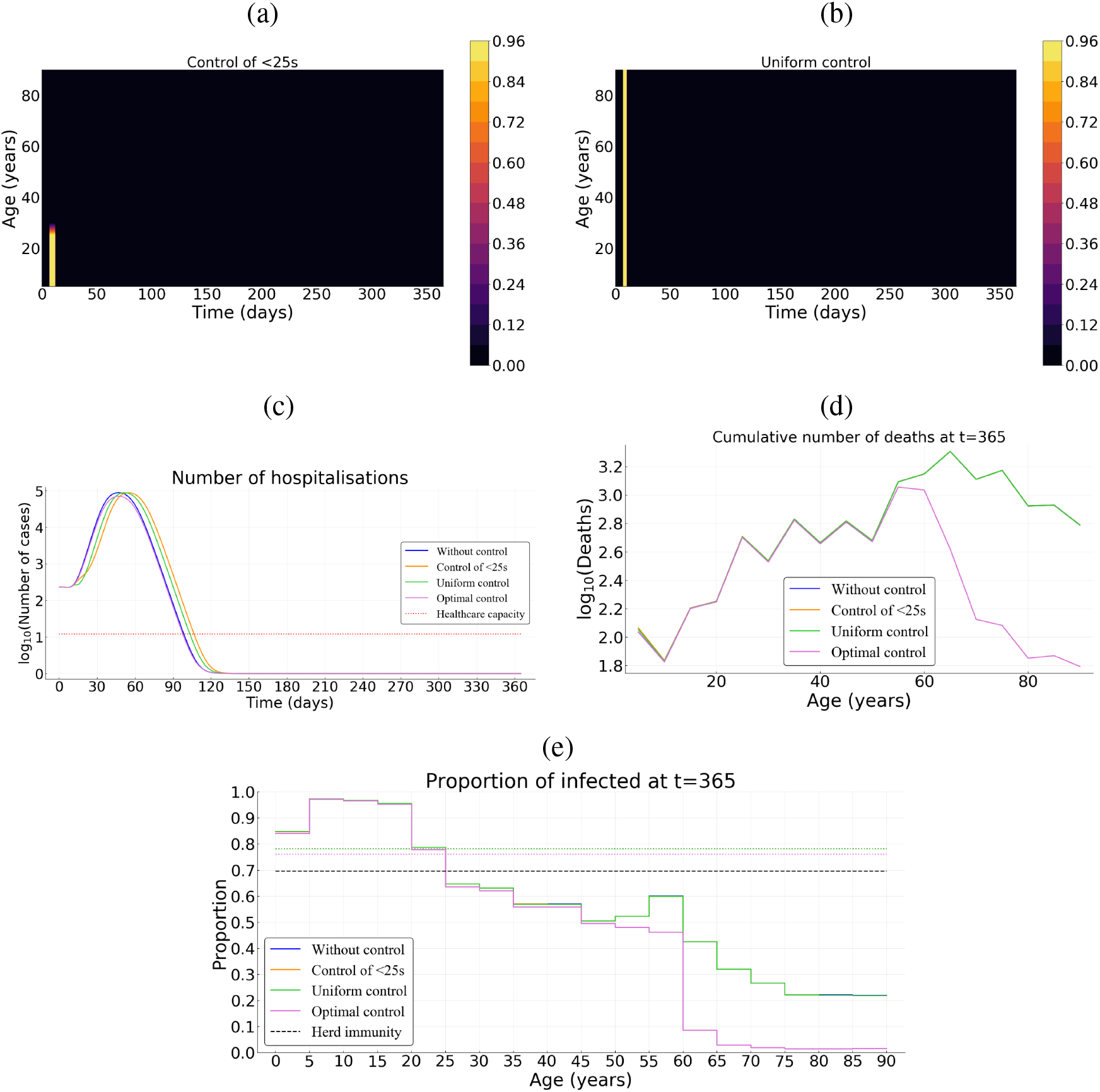
Comparing optimal control with uniform control of the whole population or over its younger fraction in Burkina Faso. (a) Illustration of the control over the young population and (b) uniform control of the whole population. (c) Number of hospitalizations. (d) Cumulative deaths per age at final time *T* = 365 days. (e) Age distribution of the proportions of the population that have been infected before one year. Here, we assume *B*\* = 10^3^ and *p* = 0.5.

A practical issue regarding the implementation of such optimal control strategy is the fact that it is a continuous function. One possibility to address this problem is to derive step functions. For instance, in Supplementary Figure S5, we subdivided the population into 10-year amplitude classes and updated the control every 3-weeks. Importantly, even though it is assumed to be constant during each 3-weeks period for each age-class, the control intensity directly originates from the results of the continuous optimal control strategy. This discrete implementation of the optimal strategy achieve similar efficiencies (Figure S5), with a relative performance of 91% compared to a doing nothing scenario.

## 6 Discussion

Non-pharmaceutical public health interventions can be implemented either to mitigate the COVID-19 epidemic wave, and rely on natural immunisation, or to suppress the wave long enough to develop and implement a vaccine or a treatment. Here, we explicitly factor in the age heterogeneity of the host population in the identification of the optimal allocation of the control efforts in three countries (Burkina Faso, France, Vietnam) which have quite contrasted age-structure of their population and social contacts (Figure 3).

We use optimal control theory to characterize an optimal strategy that significantly reduces the number of deaths, while being sustainable at the population level. Our formulation assumes a quadratic cost for the control effort. Overall, the optimal control lasts less shorter in Burkina Faso compared to cases of France and Vietnam. We find that, with this strategy, the intensity of the control is always relatively high on the older fraction of the population during at least a hundred days, before decreasing more or less rapidly depending on the cost associated to the control and the social structure of the host population. The control over the younger fraction of the population is weak and only occurs when the cost associated with the optimal control is relatively low and, while the control extends to the younger population in France and Vietnam after 2 or 3 months, such extension is very moderate (or even negligible) in Burkina Faso. This late control over the younger part of the population actually mimics the results [10] where the control did not peak right away. Intuitively, if control strategies come at a high cost for the population, it is best to focus on the age classes that are the most at risk. Conversely, if the control measures are more acceptable to the population, the optimal strategy is to aim wide in order to completely suppress the epidemic wave.

Information on the natural history of paucisymptomatic infections of COVID-19 remains relatively little-known [65, 66]. It is estimated that a proportion *p* of infected individuals will remain asymptomatic throughout the course of infection. However, this proportion remains largely unspecified in the literature [65, 66]. We explored effects of the proportion *p* on the optimal control strategy *c*\*. Overall, the proportion of paucisymptomatic infections has marginal effects on the optimal control strategy (Figure S6). The optimal control remains strong over the older population from the beginning of the epidemic, before being progressively relaxed. The control over the younger population is weaker and occurs only if the control cost itself is low. But, the level of control over the younger fraction increases when the proportion of paucisymptomatic infections decreases. Further, for high values of *B*\* (10^3^ or 10^4^), the shape of the optimal control does not change with the proportion *p* (Figure S6). Indeed, the epidemics cannot be stopped and the strategy is then to reduce mortality by protecting the population the most at risk (here the older population). However, with low value of *B*\* (10^2^), different shape of the optimal control give the same result since there are enough resources to stop the epidemics.

Given the leverage represented by school and university closure, we investigated the effect of control measures over individuals aged under 25. Our results show that NPIs targeting the younger fraction of the population are not very efficient in reducing cumulative mortality, unless they can be implemented strongly and for a relatively long period. Indeed, the number of deaths with a control only over the younger population is similar to a doing nothing scenario for cases of Burkina Faso, France and Vietnam (Figures 7, 8, S4 a, c). Thus, this result does not seems to depend on the age-structure and social contacts of the population considered. However, the variation of the transmission probability with age (discussed below) can potentially impacts a such result.

The formulation of the objective functional considered here search for the optimal control effort the cumulative number of deaths. However, other objective functions can be considered including for example, long-term hospitalizations and long-term health consequences. It is equivalent to considering the number of hospitalizations as the variable to be minimized and costs associate to long-term hospitalizations and long-term health consequences. This formulation may indeed be interesting to look at in details but would deserve to be considered independently in another study.

The model proposed here is an extension of the classical models based on ordinary differential equations that tackled the issue of the optimal control of COVID-19 outbreak [9–12]. Here, the whole population is structured by age (*a*) and additionally by the time since infection (*i*) for infectious individuals, which echoes the model developed in [64] using a discrete-time formulation of the infection. With our continuous structure, we show that the number of new cases *I*_*N*_(*t, a*) at time *t* in individuals of age *a* is given by the renewal equation

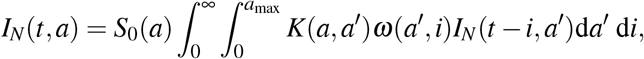

where *K* is the contact matrix and *ω*(*a, i*) is the infectiousness of individuals aged *a* which are infected since time *i* (Appendix B). For parameterisation purposes, we assume that *ω*(*a, i*) is the product between the proportion of individuals of age *a* in the whole population and the infectiousness 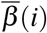 of individuals infected since time *i*. This is potentially a limitation —not in the model formulation proposed here, but rather in parameterisation perspective in relation to the existing literature— since infectiousness 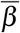 could depend on the age *a* thereby creating an additional heterogeneity in addition to that since the time since infection *i*. This issue can be particularly important since some studies suggest a low risk of transmission in the young population (*e.g*. [67]). On the other hand, although superpsreading events (of young people) have been documented, there is still much uncertainty about their relative role in the spread of the epidemic and about their origin (superspreading could be linked to environmental conditions, such as massive gatherings, rather than individual properties). Therefore, assuming independence from age seems the most parsimonious assumption given the current data.

Another potential limitation is the lack of gender structure and comorbidities in the model formulation. Given the observed male biased in mortality during the COVID-19 pandemic, it has been suggested that males are more at risk of developing severe infections [68]. This heterogeneity could readily be introduced in the model.

Contact networks have an important role in transmission dynamic models. Epidemic models that determine which interventions can successfully prevent an outbreak may benefit from accounting for social structure and mixing patterns. Contacts are highly assortative with age across a given country, but regional differences in the age-specific contacts is noticeable [43]. The current model could be modified to explore epidemiological dynamics in a spatially structured population with nonhomogeneous mixing, *e.g*. by using a meta-population model [69].

Another potential extension of the model would be to allow for the isolation of symptomatic cases and their contacts, following the method developed in [70] and applied recently to digital contact tracing [22]. Indeed, these measures strongly depend on the relative timing of infectiousness and the appearance of symptoms, and the formulation of the presented model seems suitable for that. However, this also raises technical challenges due to the double continuous structure. Being able to identify age classes to follow in priority with contact tracing could be, though, an asset in controlling epidemic spread.

## Data Availability

All data pertaining to this study are available within the manuscript

## Code availability

The code (with the Julia Programming Language) used to simulate the model can be accessed through the Zenodo platform http://doi.org/10.5281/zenodo.4288144

## A Supplementary Figures

**Figure S1:**
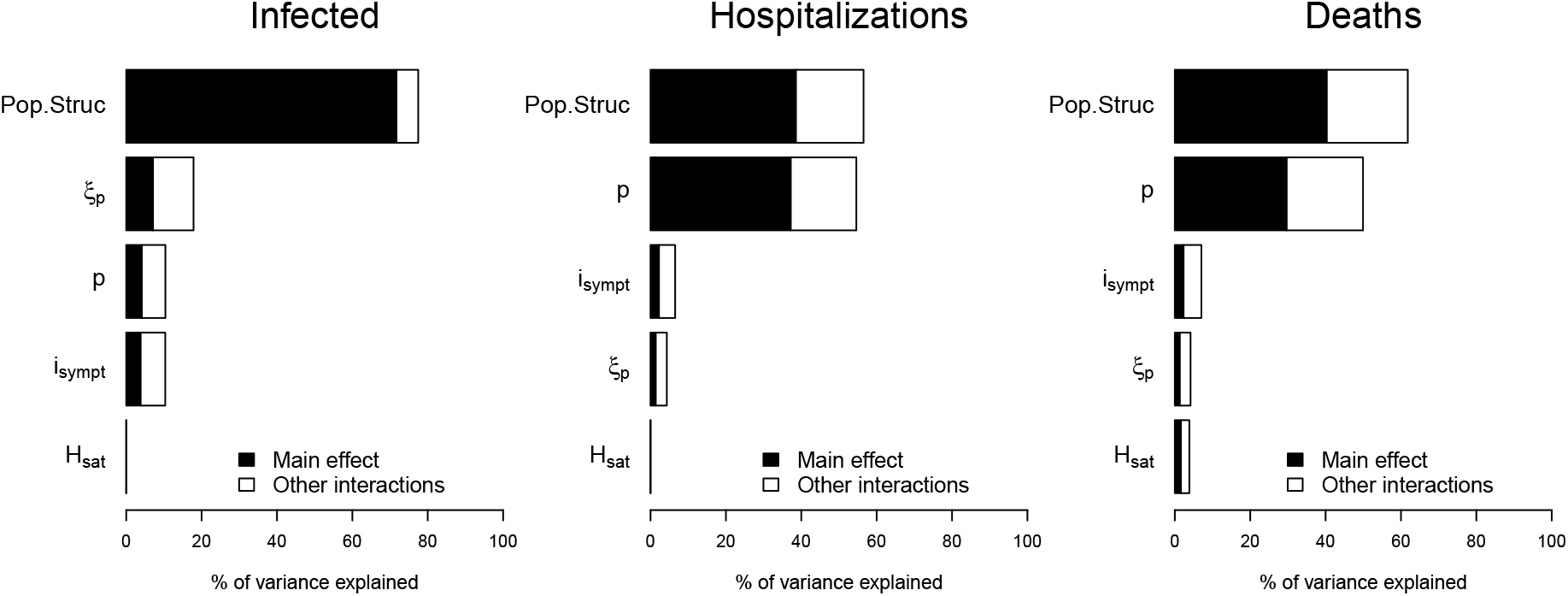
Global sensitivity analysis. Sensitivity of infected individuals, hospitalizations and deaths to the proportion of paucisymptomatic (*p*), average time of symptoms onset (*i*_*sympt*_), infectiousness reduction of paucisymptomatic (*ξ* _*p*_), healthcare capacity (*H*_*sat*_) and population structure –Pop.Struc– (including the natural mortality, the size of the population, age-structure and social contacts).

**Figure S2:**
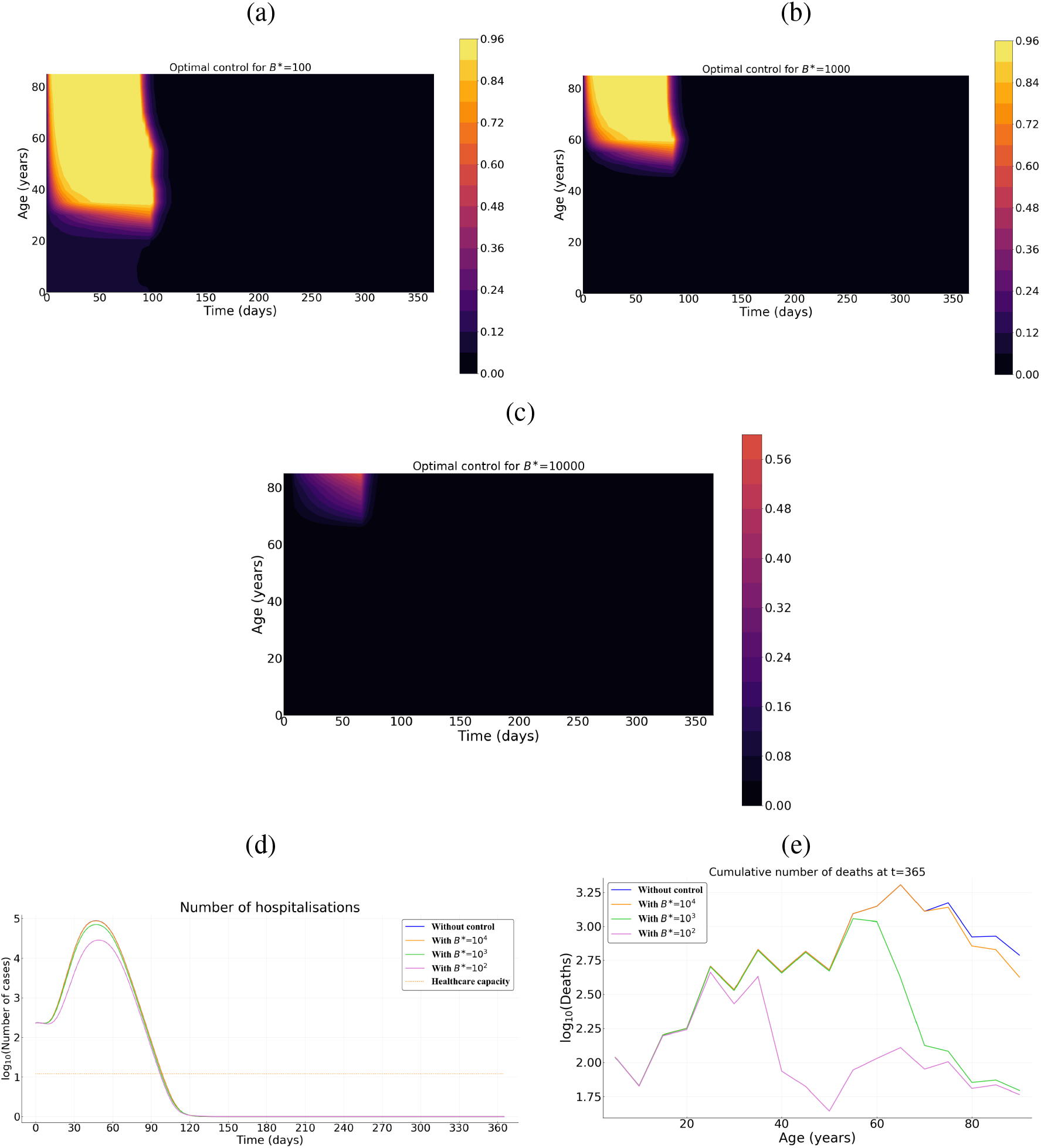
Optimal control strategy (*c*\*) as a function of the cost of the control measures in Burkina Faso. Intensity of the control as a function of time and host for for (a) relatively low *B*\* = 10^2^, (b) an intermediate *B*\* = 10^3^, and (c) a high *B*\* = 10^4^ cost. (d) Prevalence of hospitalized patients as a function of the strategy and the cost. (e) Cumulative deaths per age at the end of the time interval (when *T* = 365 days). Parameter values not related to the control are identical to Figure 4.

**Figure S3:**
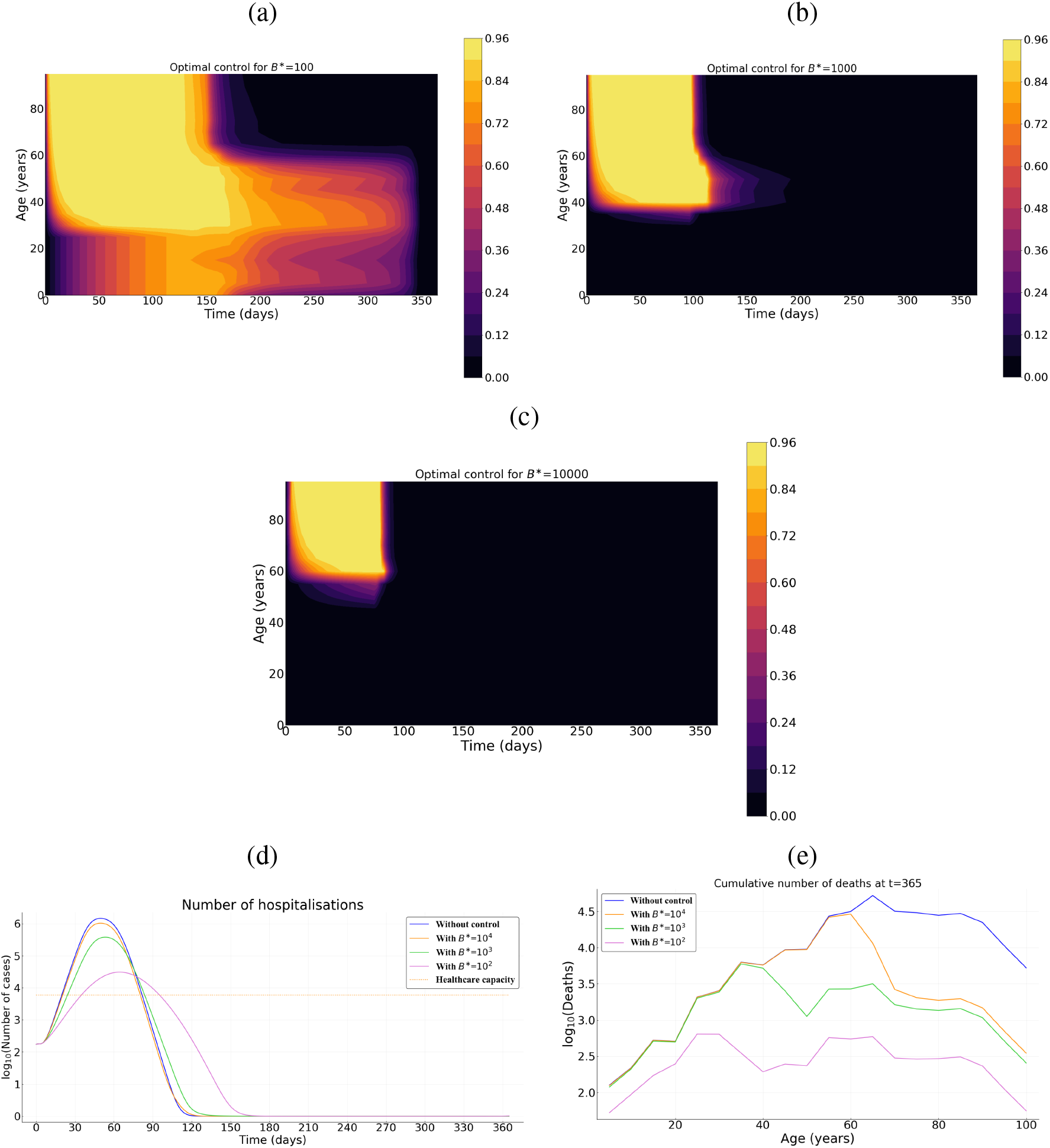
Optimal control strategy (*c*\*) as a function of the cost of the control measures in Vietnam. Intensity of the control as a function of time and host for for (a) relatively low *B*\* = 10^2^, (b) an intermediate *B*\* = 10^3^, and (c) a high *B*\* = 10^4^ cost. (d) Prevalence of hospitalized patients as a function of the strategy and the cost. (e) Cumulative deaths per age at the end of the time interval (when *T* = 365 days). Parameter values not related to the control are identical to Figure 4.

**Figure S4:**
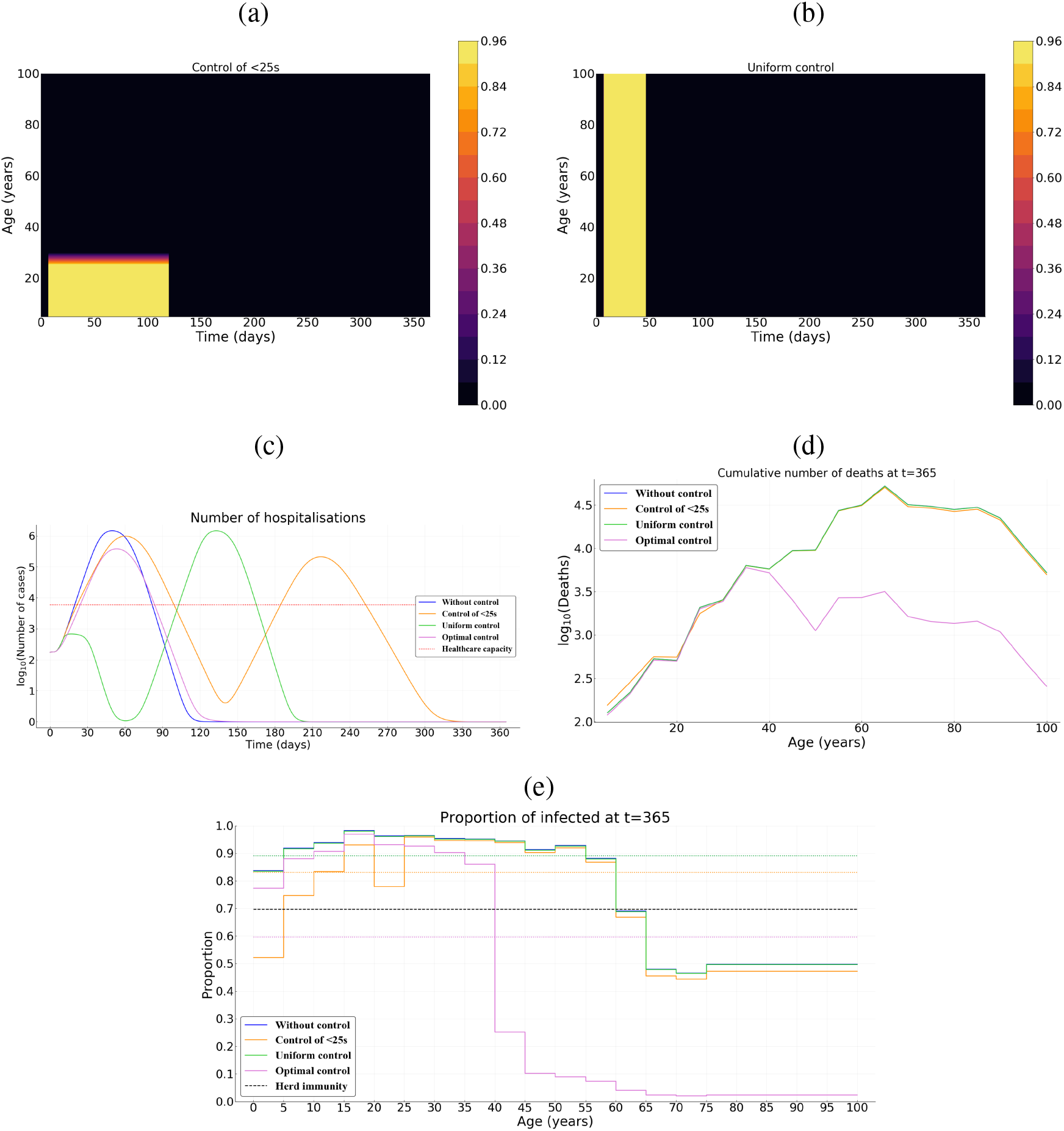
Comparing optimal control with uniform control of the whole population or over its younger fraction in Vietnam. (a) Illustration of the control over the young population and (b) uniform control of the whole population. (c) Number of hospitalizations. (d) Cumulative deaths per age at final time *T* = 365 days. (e) Age distribution of the proportions of the population that have been infected before one year. Here, we assume *B*\* = 10^3^ and *p* = 0.5.

**Figure S5:**
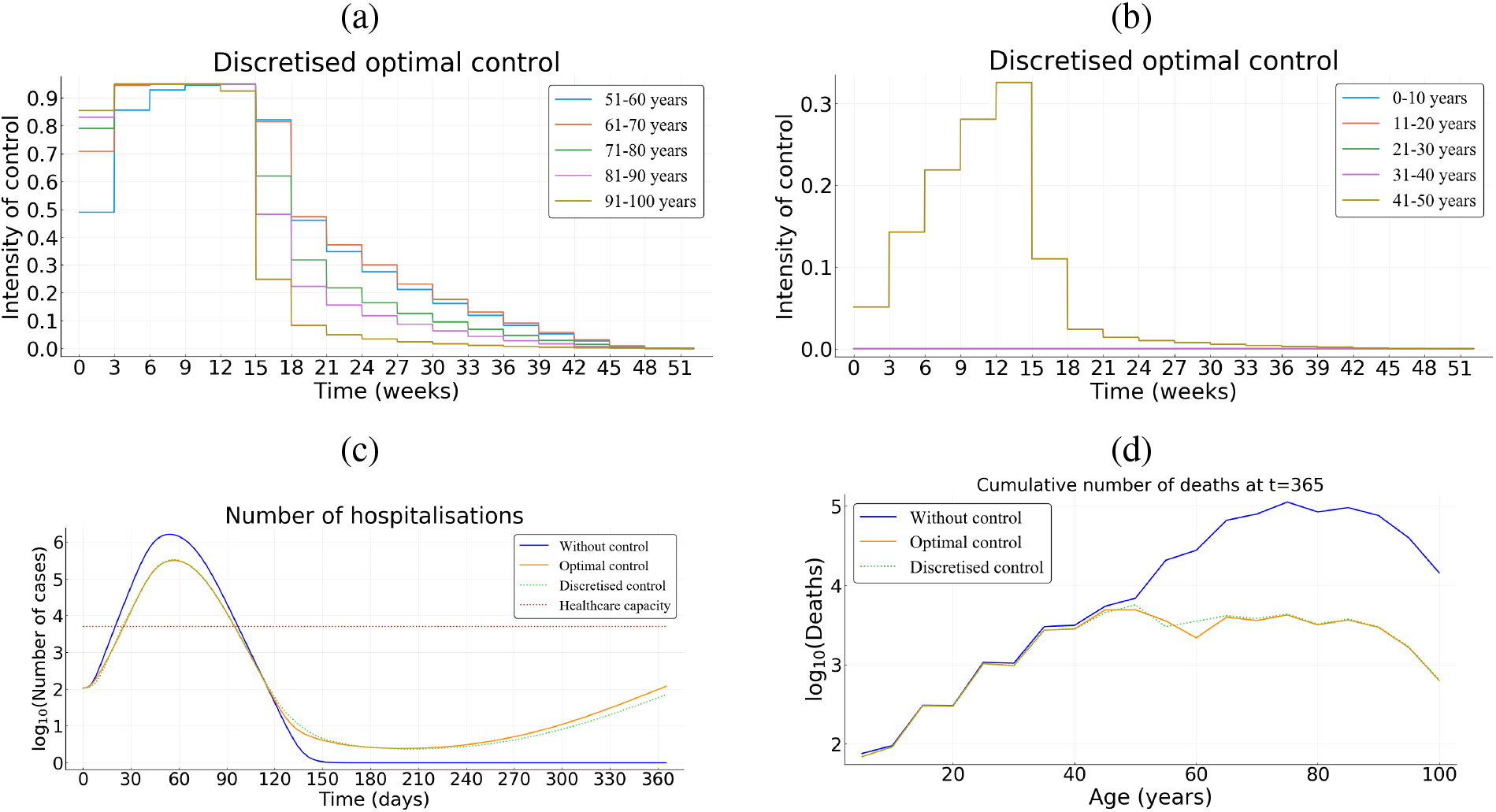
Practicability of the age-structured optimal control. (a)-(b) Step optimal controls with a 3-weeks update over the older and younger populations. The corresponding optimal is given by Figure 6 b. (c)-(d) Cumulative deaths per age at final time *T* = 365 days.

## B Basic reproduction number

Here we compute the basic reproduction number *R*_0_ of the model (3)-(5). First let us set for *i* ≥ 0 and *a* ∈ [0, *a*_max_] the following functions

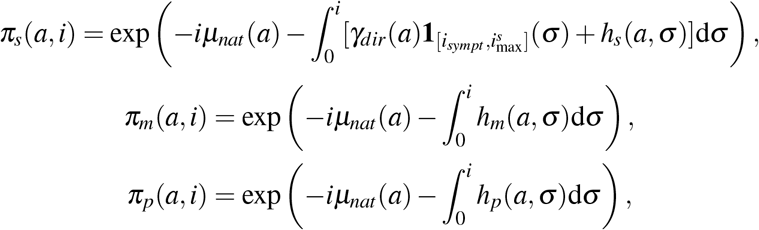

that describe the survival probability of infected individuals (in the respective compartment), with age *a*, from their infection until the time since infection *i*, in case of no hospitalisation (*i.e. H* ≡ 0). We get the following Volterra formulation of the linearized system of (3)-(5):

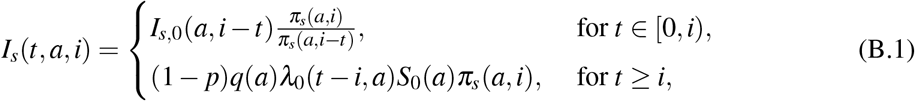

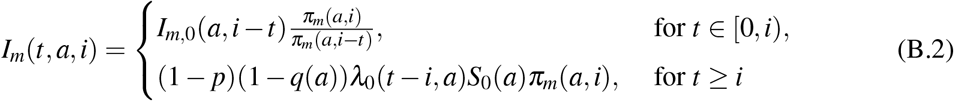

**Figure S6:**
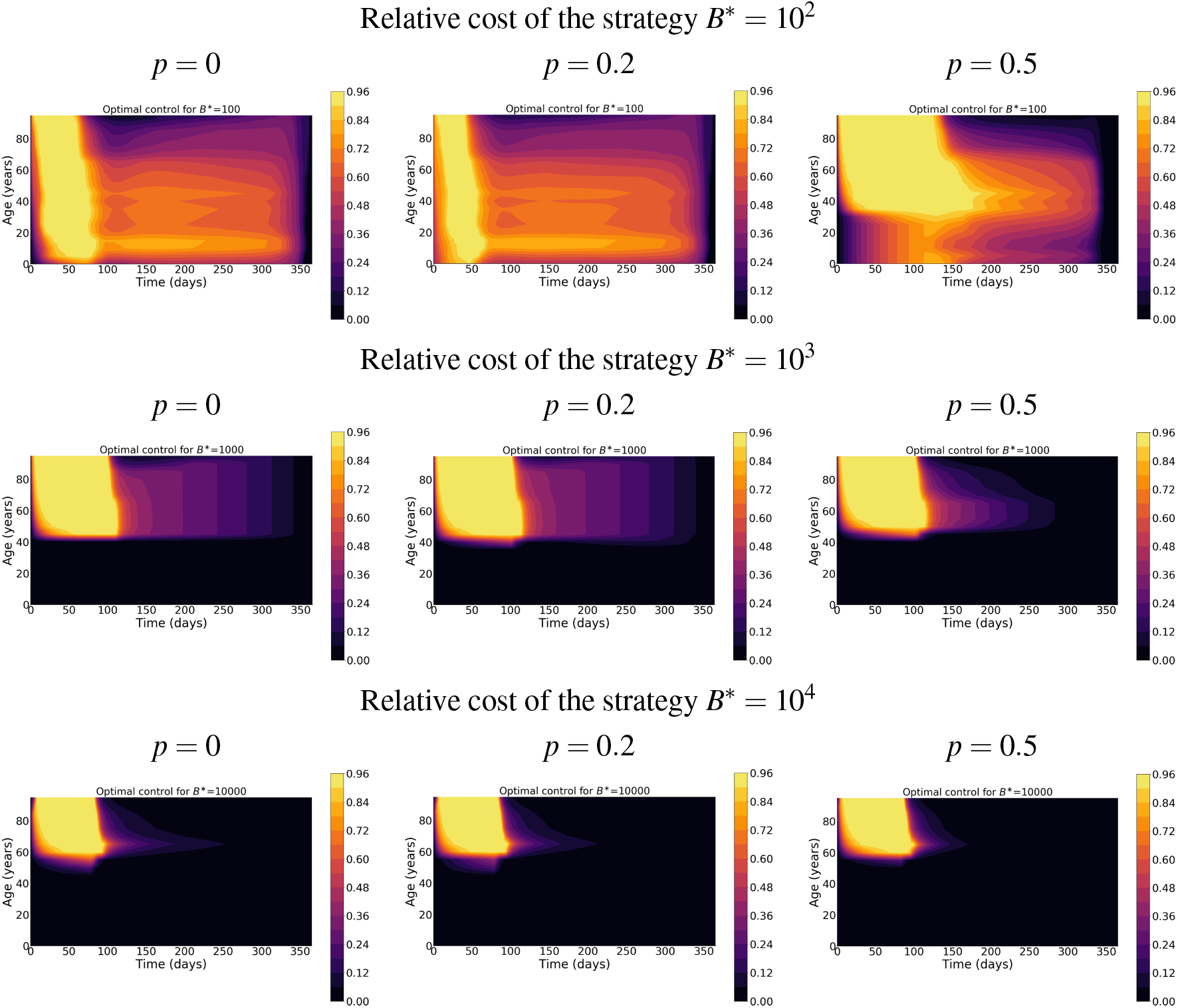
The effect of paucisymptomatic infections, through their proportion *p*, on the optimal control *c*\*.

and

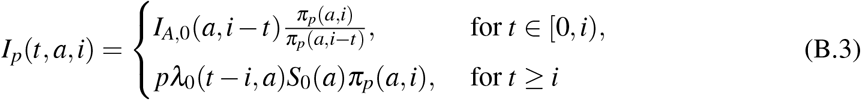

where *λ*_0_ = *λ* (·, ·, 0) is defined by

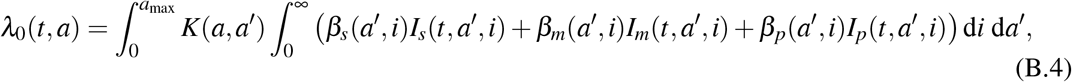

where *β*_*k*_, *k* ∈ {*s, m, p*} are defined in Section 3.2. Let *I*_*N*_(*t, a*) = *λ*_0_(*t, a*)*S*_0_(*a*) be the density of newly infected of age *a* at time *t*, with *c* ≡ 0. Then (B.1)-(B.2)-(B.3) can be rewritten as the following Volterra formulation:

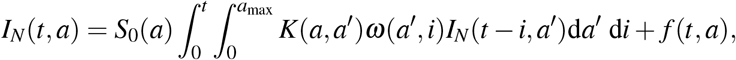

where

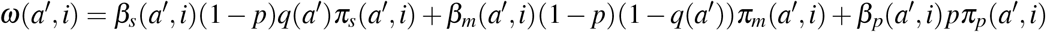

and *f* (*t, a*) is the density of new infections produced by the initial population. Therefore, the basic reproduction number *R*_0_ is the spectral radius, denoted by *r*(*U*), of the next generation operator *U* defined on 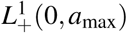 by

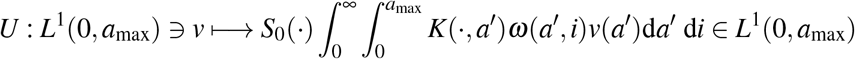

As explained in Section 3.2, it is estimated in [22] that each average infectiousness *β*_*k*_ (*k* ∈ {*s, m, p*}) takes the form of a Weibull distribution *W* (3, 5.65) so that the mean and median are equal to 5.0 days while the standard deviation is 1.9 days. Based on this estimation, we assume that 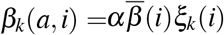 where 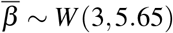 and *α* is a positive parameter to be determined. Consequently, it follows that *α* is given by

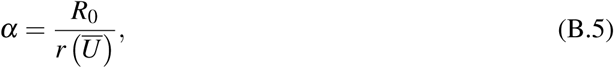

where *Ū* is the operator defined by

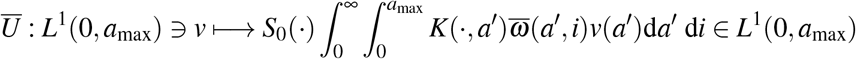

with

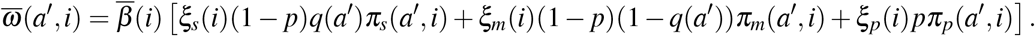

We see that *Ū* can be rewritten as

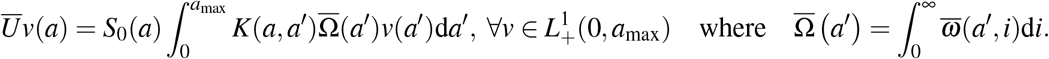

Now, in order to compute the spectral radius *r*(*Ū*), we first make the following assumptions: **Assumption B.1** *We suppose that functions S*_0_,*K*, 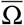 *are bounded and positive almost everywhere*. Then, we can show that r*(Ū*) is given by the spectral radius of the following linear operator:

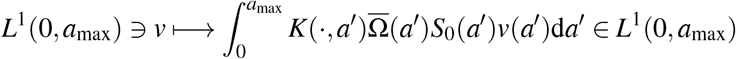

which can be easily computed since the age *a* is numerically divided into *N* classes, so that the term inside the integral of the latter equation is a *N* × *N* matrix. Finally, we obtain *α* from (B.5).

In addition to Assumption B.1, if the function *K* is symmetric, we can define the positive selfadjoint operator *S* by

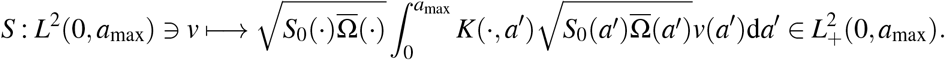

We then deduce the following

### Proposition B.2

*Let K be symmetric and Assumption B.1 be satisfied. Then, operators U and S are positive and compact, their spectra σ(Ū*) \ {0} *and σ* (*S*) \ {0} *are composed of isolated eigenvalues with finite algebraic multiplicity. Moreover, we have σ (Ū*) = *σ(S) ⊂ ℝ* _*+*_ *and the following Rayleigh formula holds:*

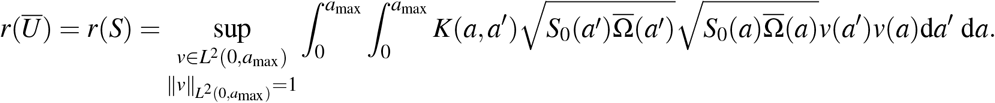

*Proof*. The compactness of both integral operators follows from the fact that *a*_max_ *<* ∞ by assumption (see Table 1), hence their spectra are punctual. Now we prove that *σ* (*Ū*).= *σ*(*S*) Let *𝒱* ∈ *σ* (*Ū*) be an eigenvalue of *Ū* and *ϕ* ∈ *L*^1^(0, *a*_max_) be the associated eigenvector, *i.e*.

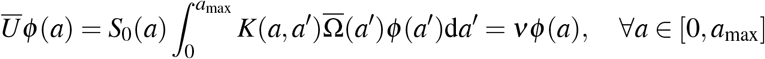

so that *ϕ* ∈ *L*^∞^(0, *a*_max_) ⊂ *L*^2^(0, *a*_max_). Defining the function

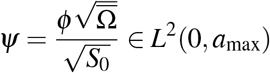

leads to

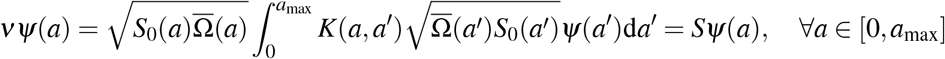

*i.e. 𝒱* ∈ *σ* (*S*) is an eigenvalue of *S* associated to the eigenvector *Ψ*, so that *σ*(*Ū*); *⊂ σ*(*S*.). For the reverse inclusion, let *𝒱* ∈ *σ* (*S*) and *Ψ* ∈ *L*^2^(0, *a*_max_) ⊂ *L*^1^(0, *a*_max_) be the associated eigenvector for *S*. It follows that the function

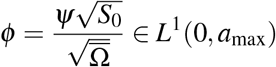

is an eigenvector of *Ū* related to the eigenvalue *𝒱* ∈ *σ* (*Ū)*, whence *σ* (*Ū)* = *σ(S)*. In particular, both spectral radius are equal. Finally, the Rayleigh formula is classical for positive and symmetric operators.

## C Computations of the adjoint system

In order to deal with the necessary optimality conditions, we use some results in [61]. Next, we detail the computations of the adjoint system (12)-(13). To this end, we first define the functions *y*_1_, *Q* : [0, *T*] × [0, *a*_max_] → ℝ and *y*_2_ : [0, *T*] × [0, *a*_max_] × ℝ_+_ by:

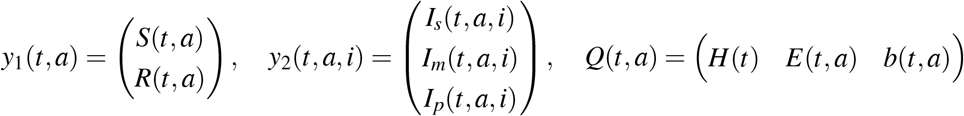

Wherein

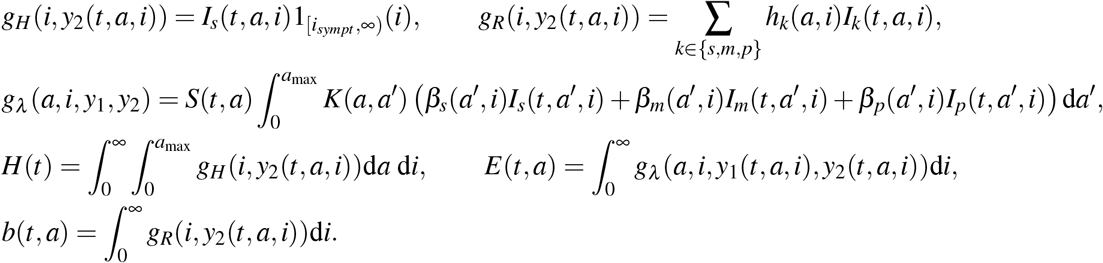

The model (5) thus rewrites as

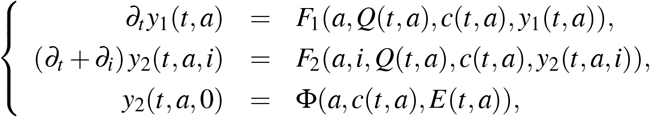

with

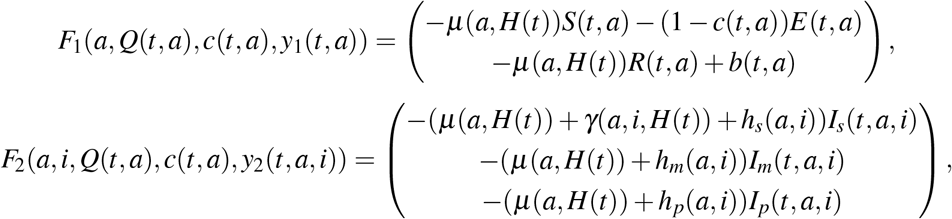

and

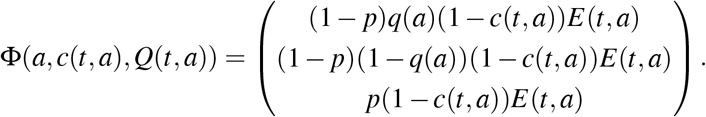

We now rewrite the functional *J* as

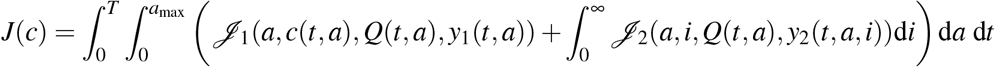

which is decomposed into

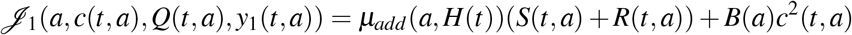

and

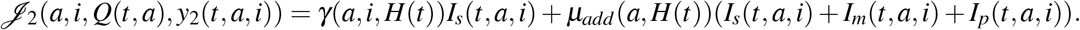

We denote by *z*_1_, *ζ* _*k*_ : [0, *T*] × [0, *a*_max_] → ℝ (for *k* ∈ {1, 2, 3}) the following adjoint functions

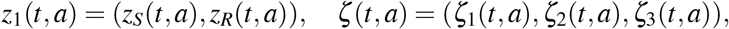

and we denote by *z*_2_ : [0, *T*] × [0, *a*_max_] × ℝ_+_ the following adjoint function

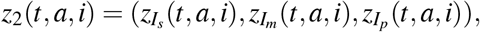

satisfying lim_*i*→∞_ *z*_2_(*t, a, i*) = 0 and *z*_1_(*T, a*) = *z*_2_(*T, a, i*) = 0. We get

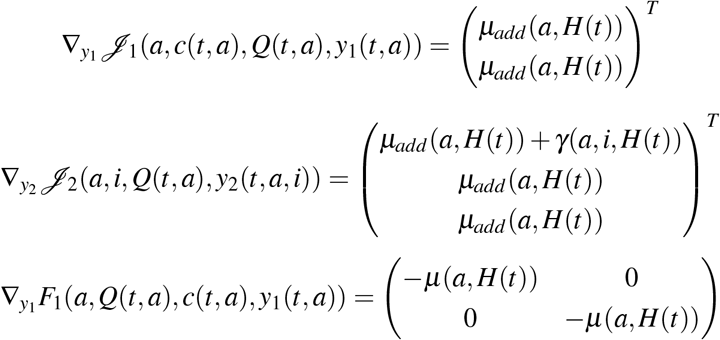

and

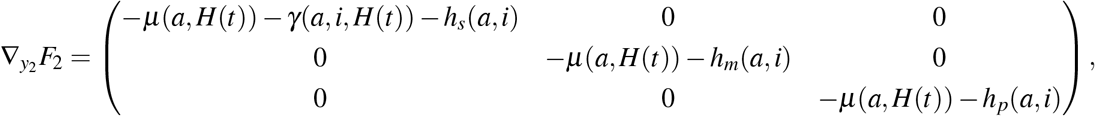

where ∇_*y*_*F* denotes differentiation of *F* with respect to the variable *y*.

Then

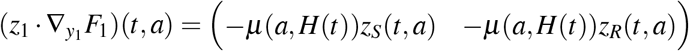

and

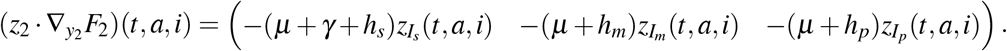

Setting

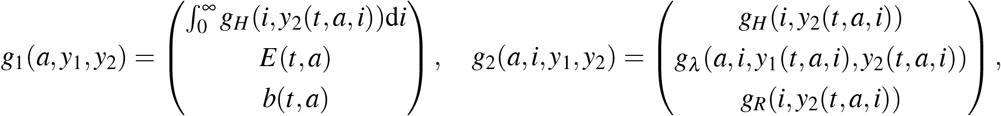

we see that

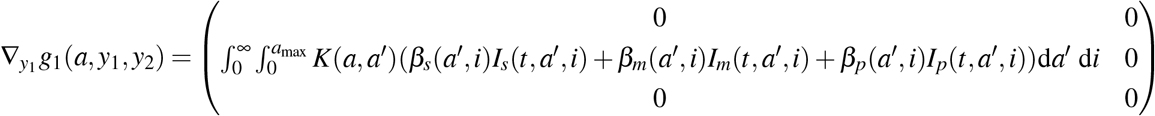

and

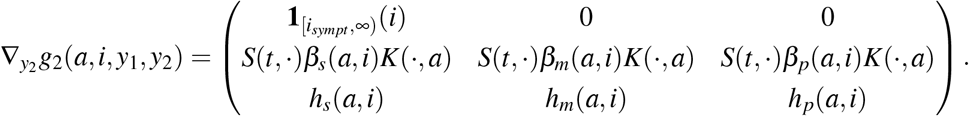

From there, we deduce that

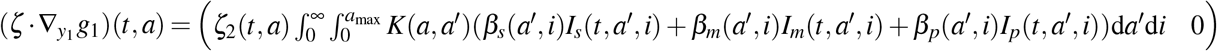

and

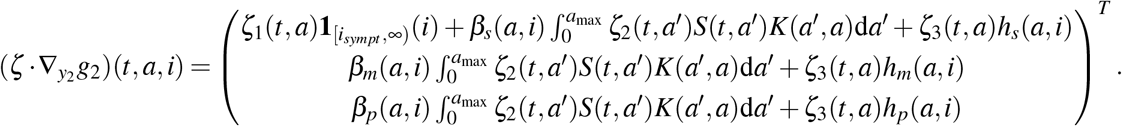

The adjoint system is given by

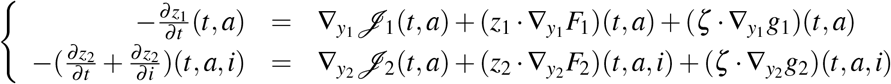

which is equivalent to (12). Next, we see that

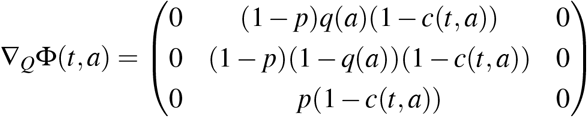

Whence

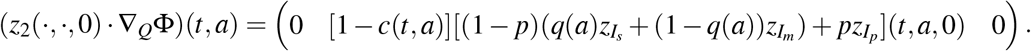

Further, we have

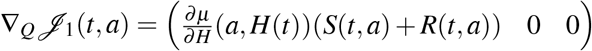

and

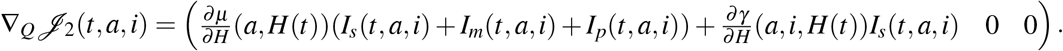

We also see that ∇_*Q*_*g*_1_ ≡ 0, ∇_*Q*_*g*_2_ ≡ 0,

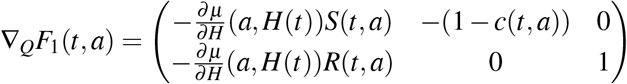

and

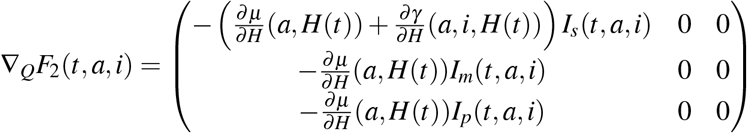

whence

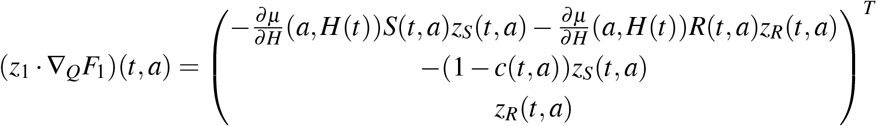

and

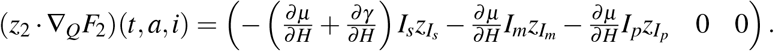

Finally, the adjoint functions *ζ* must satisfy the following equation:

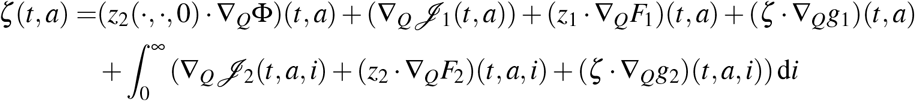

which is equivalent to (13). Finally by [61], the Hamiltonian is given by

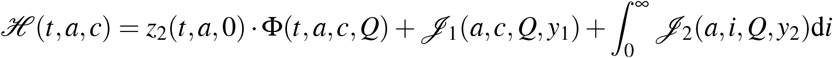

which leads to

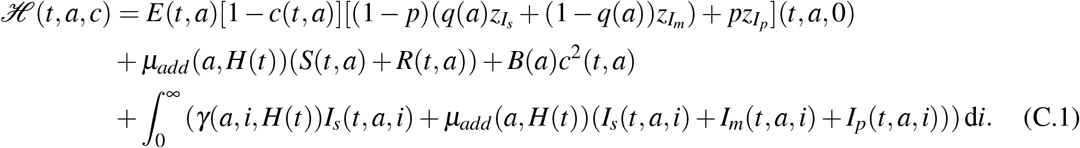

## Notes

### Competing Interest Statement

The authors have declared no competing interest.

### Funding Statement

No external funding was received

### Summary of Updates

In particular, we now apply our age-structured model to three countries with contrasted demography: France, Vietnam, and Burkina Faso. Generating more concrete results allows us to better illustrate the potential of the model.

